# Sound Exposure During Sleep (SES) in PTSD Patients: An Open-Label Feasibility Study

**DOI:** 10.64898/2026.05.02.26352243

**Authors:** Keiko Ino, Keiichi Zempo, Arinobu Hori, Tatsuya Maruyama, Morie Tominaga, Yuki Sugaya, Mari Oba, Yuta Yamauchi, Luna Sato, Haruka Sekiba, Chinatsu Kawakami, Gabriel Bachman, Izumi Waki, Hiroyuki Kitagawa, Masashi Yanagisawa, Yoshiharu Kim, Masanori Sakaguchi

**Affiliations:** Department of Behavioral Medicine, National Institute of Mental Health, National Center of Neurology and Psychiatry (NCNP), Kodaira, Tokyo, Japan; Institute of Systems and Information Engineering, University of Tsukuba, Tsukuba, Ibaraki, Japan; Hori Mental Clinic, Fukushima, Japan; Clinical Research Promotion Centre, The University of Tokyo Hospital, Tokyo, Japan; S’UIMIN Inc., Tokyo, Japan; Department of Neurophysiology, Graduate School of Medicine, The University of Tokyo, Tokyo, Japan; International Research Center for Neurointelligence (WPI-IRCN), The University of Tokyo Institutes for Advanced Study (UTIAS), Tokyo, Japan; Department of Clinical Data Science, Clinical Research and Education Promotion Division, National Center Hospital, National Center of Neurology and Psychiatry (NCNP), Kodaira, Tokyo, Japan; Graduate School of Systems and Information Engineering, University of Tsukuba, Tsukuba, Ibaraki, Japan; Tsukuba Institute for Advanced Research (TIAR) and International Institute for Integrative Sleep Medicine (WPI-IIIS), University of Tsukuba, Tsukuba, Ibaraki, Japan; Master’s Program in Medical Science, Graduate School of Comprehensive Human Science, University of Tsukuba, Tsukuba, Ibaraki, Japan; Ph.D. Program in Humanics, Graduate School, University of Tsukuba, Tsukuba, Ibaraki, Japan; Tsukuba Clinical Research and Development Organization (T-CReDO), University of Tsukuba, Tsukuba, Ibaraki, Japan; University of Texas Southwestern Medical Center, Dallas, TX, USA

## Abstract

Trauma-focused psychotherapies for post-traumatic stress disorder (PTSD) require waking re-engagement with traumatic memories, driving high dropout. We tested whether trauma-linked auditory cues delivered during slow-wave sleep are feasible. Of 13 patients who provided written informed consent, 6 (100% female) completed overnight Sound Exposure during Sleep (SES); none of the adverse events observed during overnight stimulation were judged by the study team to be attributable to the auditory intervention, and slow-wave sleep was preserved. Two sequential protocol versions were used: Version A (n = 2; capped at SUDs 30–40) and a no-ceiling amendment (Version B, n = 4). Post-hoc exploratory analyses (not powered for efficacy) showed Version B reduced subjective distress (mean difference −65.5%, 95% CI −104.2 to −26.7; nominal p = 0.012) and PCL-5 intrusion (−7.0; nominal p = 0.015). Findings are exploratory and require sham-controlled confirmation. Trial registration: jRCT1030230706.

## Introduction

Post-traumatic stress disorder (PTSD), as defined by the Diagnostic and Statistical Manual of Mental Disorders, Fifth Edition (DSM-5), develops following exposure to actual or threatened death, serious injury, or sexual violence and is characterized by intrusive re-experiencing, persistent avoidance of trauma-related stimuli, negative alterations in cognition and mood, and marked alterations in arousal and reactivity [1]. In the United States, approximately 5% of adults—roughly 13 million individuals—meet criteria for PTSD in a given year, and lifetime prevalence approaches 6–7% [2]. Globally, lifetime prevalence is estimated at approximately 3.9%, with an estimated 13 million new cases emerging annually [3,4]. Despite this burden, treatment engagement remains limited: approximately 50% of individuals with PTSD in high-income countries receive any form of treatment, compared to fewer than 20% in low- and middle-income countries [3]. Given the substantial personal, societal, and economic impact of PTSD—including an estimated $232.2 billion in annual healthcare and productivity costs—there remains a critical need for effective, scalable, and accessible interventions [5].

Current clinical guidelines recommend trauma-focused psychotherapies as first-line treatments for PTSD and identify the selective serotonin reuptake inhibitors (SSRIs) sertraline and paroxetine as the only FDA-approved pharmacologic options for PTSD [6]. The 2023 VA/DoD Clinical Practice Guideline additionally endorses the serotonin–norepinephrine reuptake inhibitor venlafaxine extended-release as a first-line pharmacotherapy, though it is not FDA-approved for PTSD; no new agents have been FDA-approved for PTSD since paroxetine (2001), with recent applications for brexpiprazole-plus-sertraline and MDMA-assisted therapy both receiving Complete Response Letters in 2024–2025.

A recent systematic review and meta-analysis of 34 randomized clinical trials (n = 3,208) found that loss of PTSD diagnosis after individual trauma-focused psychotherapies ranged from 65% to 86% in civilian samples and 44% to 50% in military and veteran samples, underscoring that remission remains far from universal and is particularly limited among veterans [7]. Sertraline and paroxetine show only modest efficacy (effect size [ES] = 0.23; 21 studies, n = 3,932), with 20–30% of patients failing to respond [8]. Side effects—such as sexual dysfunction, weight gain, and sleep disruption—limit tolerability and contribute to a 9% dropout rate [9–11]. Dropout also remains a major challenge in PTSD psychotherapy, averaging 16% across 116 studies and rising to 24.2% among veterans (n = 2,984) [12,13]; contributing factors include logistical barriers and therapy-induced distress [14]. Even among those who initially remit, about 13.1% relapse (21 studies) [15], often due to ongoing stress, treatment discontinuation, or residual symptoms. These issues underscore the need for more flexible, patient-centered interventions—such as home-based or remote options—and sustained long-term support to reduce relapse.

A range of novel pharmacotherapies and drug-assisted interventions are currently under investigation to address the limitations of existing PTSD treatments. The most prominent recent efforts have centered on MDMA-assisted therapy; despite demonstrating substantial symptom reductions in two randomized Phase 3 trials [16,17], the U.S. Food and Drug Administration (FDA) declined immediate approval in 2024, citing concerns over functional unblinding, expectancy bias, adverse-event reporting, and durability, and requested additional trial data [18]. In parallel, ketamine has garnered significant attention, with randomized trials showing rapid but largely transient reductions in PTSD severity following repeated intravenous infusions [19]; current clinical practice guidelines, however, recommend against its routine use for PTSD [6,20]. Atypical antipsychotics (e.g., risperidone, quetiapine) have historically seen extensive off-label prescription, but the 2023 VA/DoD Clinical Practice Guideline recommends against their routine use for core PTSD symptoms owing to limited efficacy and substantial metabolic adverse-effect burdens [6]. As pharmacological approaches continue to face regulatory and safety hurdles, psychological and sleep-based therapies remain critical complementary strategies.

Among first-line trauma-focused psychotherapies, Prolonged Exposure (PE)—which weakens trauma-related fear through repeated, controlled re-engagement with trauma cues, a learning process termed fear extinction—generally outperforms pharmacologic options. In randomized trials comparing PE and SSRI treatment over two years, PE more often led to loss of PTSD diagnosis (number needed to treat for an additional beneficial outcome [NNTB] = 7.0), clinically meaningful symptom improvement (NNTB = 5.7), and greater reductions in self-reported PTSD (ES = 0.39), depression (ES = 0.37), and anxiety (ES = 0.44; n = 200) [21]. Despite this superiority over medication, fear-extinction-based therapies are demanding; PE, for example, typically requires 12–16 weekly 90-minute sessions plus daily homework, and such protocols are experienced as psychologically stressful because they require repeated conscious confrontation with trauma-related cues [22]. These practical and emotional burdens—which contribute to high dropout rates and limit treatment access [12–14]—motivate interest in approaches that can modify fear memories without requiring the patient to consciously re-engage with trauma during wakefulness. Because fear-extinction memory consolidation is modulated by sleep—including slow-wave and REM sleep stages—with accumulating evidence that this sleep–extinction relationship may be altered in PTSD and related trauma disorders [23,24], targeted modulation of trauma memory during sleep represents a biologically plausible intervention window worthy of empirical investigation.

One such approach is Targeted Memory Reactivation (TMR), which presents sensory cues—such as sounds—during sleep to selectively reactivate, and thereby modulate, specific memories previously associated with those cues during wakefulness [25]. TMR during slow-wave sleep (SWS) has been shown to influence emotional memory processing [26,27]. A recent sleep-based approach augmenting Eye Movement Desensitization and Reprocessing (EMDR) therapy (hereafter, EMDR-augmented TMR) reported preliminary efficacy in reducing PTSD symptoms but had notable limitations: the TMR and sham groups differed at baseline in avoidance scores, and group differences on most clinical outcomes—including overall PTSD symptom severity—did not reach statistical significance [28]. A complementary approach in patients with nightmare disorder demonstrated that delivering TMR cues during REM sleep—specifically utilizing sounds previously paired with positive waking imagery rehearsal therapy—resulted in sustained reductions in nightmare frequency and distress [29]. Relatedly, a recent study extended REM-sleep TMR to augment imagery rescripting of negative autobiographical memories in healthy volunteers, demonstrating that targeted reactivation during sleep can consolidate the emotional modulation achieved by waking therapy [30]. In healthy human models, playing fear-conditioned tones during SWS has successfully facilitated fear extinction without waking exposure in both healthy humans [31,32] and rodent models [33], with a recent meta-analysis supporting generalizability across TMR paradigms [34]. However, crucially, in these prior paradigms the reactivated cues were experimentally conditioned to aversive stimuli, or were signals associated with a concurrent waking psychotherapy, rather than cues intrinsically tied to each participant’s lived traumatic event. No human study, to our knowledge, has yet reactivated the autobiographical trauma memory itself during sleep as a stand-alone intervention.

By contrast, the present study delivers auditory stimuli intrinsically tied to each participant’s autobiographical trauma memory during sleep, aiming to test whether sleep-targeted delivery of trauma-linked cues is feasible while exploring possible memory modulation. We conducted a single-arm, open-label feasibility trial of Sound Exposure during Sleep (SES). To our knowledge, SES is the first human intervention to deliver autobiographical trauma cues during sleep as a stand-alone approach, reducing the need for concurrent waking psychotherapy. By delivering each participant’s own personalized cues during SWS, SES is designed as a candidate intervention that may facilitate modulation of trauma memory representations without requiring deliberate, therapeutic waking re-engagement with the traumatic event.

## Results

### Participant Flow and Baseline Characteristics

The full study protocol is shown in Figure 1. Briefly, the protocol comprised eligibility screening and baseline assessments (Visit 0), creation and waking validation of a personalized autobiographical trauma-linked auditory stimulus (Visits 0–3), the overnight SES intervention at the Phase 1 (P1) Unit (Visit 4)—a brief pre-sleep waking exposure to the finalized sound followed by repeated presentation of the same stimulus during slow-wave sleep (SWS)—and a follow-up assessment within 1–4 weeks post-intervention (Visit 5). The primary endpoint was procedural feasibility; safety was assessed throughout, with secondary exploratory endpoints including subjective distress (SUDs), PTSD symptom severity (PCL-5, PDS-5), and physiological measures (HRV, SCL) during sound stimulation.

**Figure 1.**
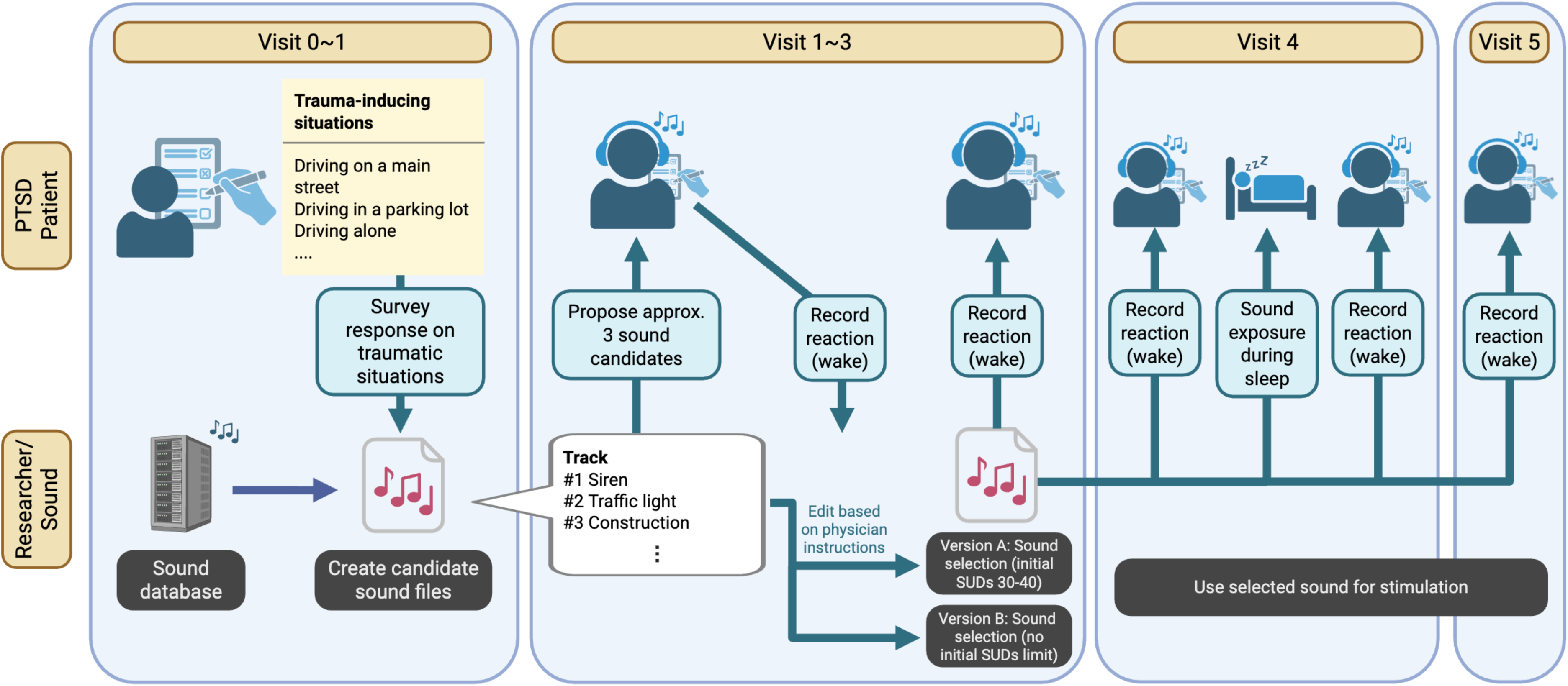
Study protocol overview. For each enrolled participant, personalized auditory stimuli reminiscent of contexts associated with the individual’s reported trauma history were developed in collaboration with treating clinicians. Trauma-context labels and sound-type icons shown in the figure are illustrative examples only; they do not represent the actual content of any single participant’s stimulus. Specific per-participant sound descriptions are not disclosed publicly to minimize re-traumatization risk in trauma-affected readers; details are available to qualified researchers upon reasonable request, subject to ethics committee approval. Created in BioRender. Sato, L. (2026) https://BioRender.com/z3z5ovp.

The flow of patient selection is shown in Figure 2. A total of 13 patients provided written informed consent between September 2024 and June 2025. Five patients were excluded prior to sound creation because of a low Posttraumatic Diagnostic Score (n = 1) or other criteria including short SWS duration (n = 4). The remaining eight patients entered the sound-development phase, during which candidate personalized trauma-associated stimuli were prepared. Of these eight, two were subsequently excluded before a final SES stimulation sound could be finalized for overnight delivery: one (H06) was deemed ineligible at the confirmatory recording due to insufficient SWS duration, and one (H05) dropped out prior to the Phase 1 (P1) Unit overnight stimulation (visit 4). Thus, a finalized SES stimulation sound was reached and delivered in six patients (visit 4). All six patients also attended the scheduled follow-up visit (visit 5) within 1 to 4 weeks after stimulation.

**Figure 2.**
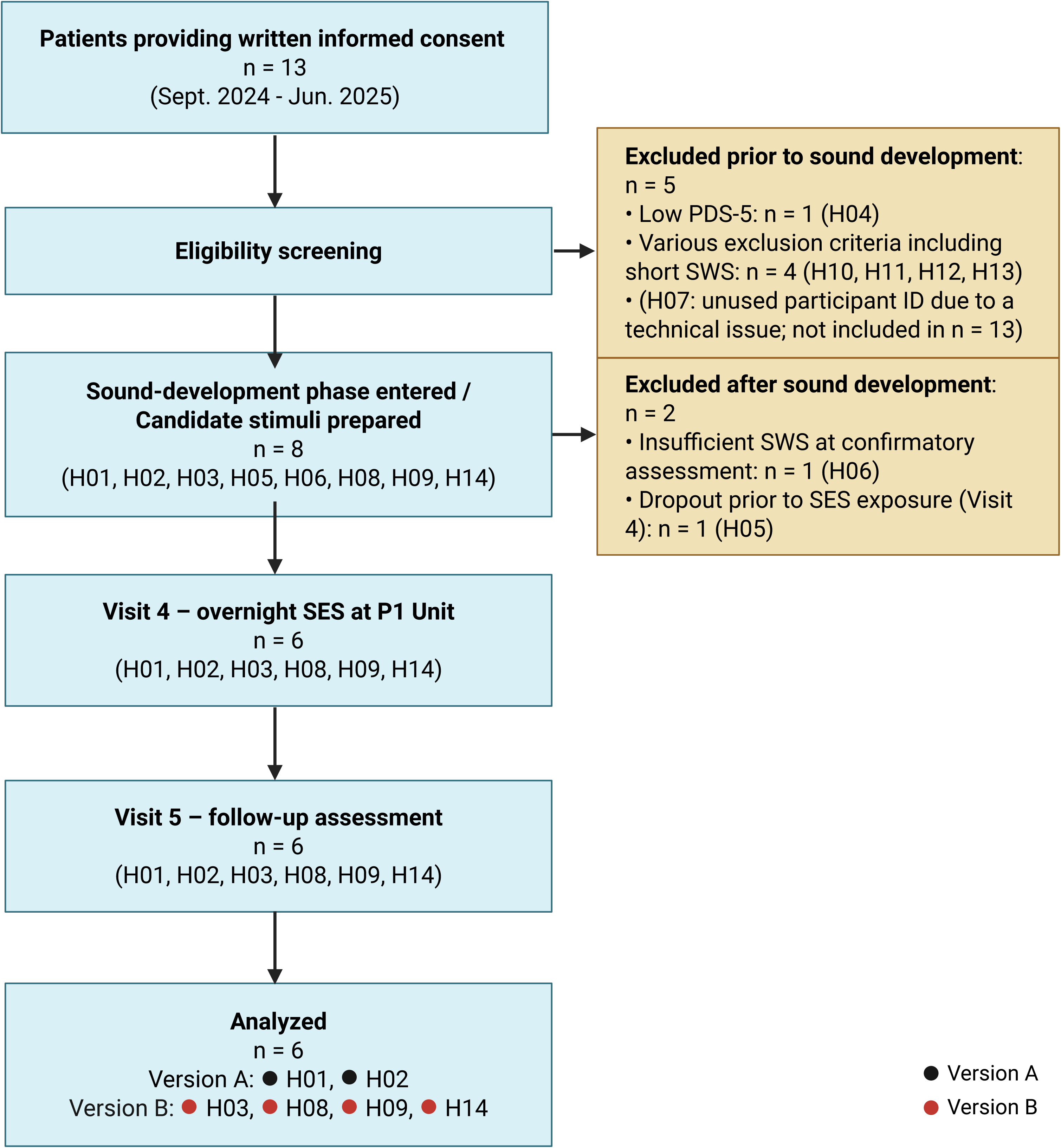
CONSORT flow diagram. A total of 13 patients provided written informed consent between September 2024 and June 2025. Five patients were excluded prior to sound creation because of a low Posttraumatic Diagnostic Score (n = 1; H04) or other criteria including short SWS duration (n = 4; H10, H11, H12, H13); H07 was an unused identifier due to a technical issue and is not counted in n = 13. The remaining eight patients (H01, H02, H03, H05, H06, H08, H09, H14) entered the sound-development phase with candidate personalized stimuli prepared. Of these eight, two were subsequently excluded before a final SES stimulation sound could be finalized for overnight delivery: one (H06) was deemed ineligible at the confirmatory recording due to insufficient SWS duration, and one (H05) dropped out prior to the P1 Unit overnight stimulation (visit 4). Thus, sound stimulation was completed in six patients (Version A: H01, H02; Version B: H03, H08, H09, H14). All six patients also attended the scheduled follow-up visit (visit 5) within 1 to 4 weeks after stimulation. Created in BioRender. Sato, L. (2026) https://BioRender.com/rqf3w40.

Notably, all six participants who completed the overnight SES procedure had successfully finalized their personalized auditory stimulus by Visit 2, without requiring an additional Visit 3 sound-development session.

Patient characteristics are presented in Table 1. All six participants were female, with a mean age of 40.5 years (SD = 8.3). One patient was in her 20s (16.7%), and two in their 30s (33.3%). The remaining three participants were above 40 years of age (50%). STOP-BANG, an eight-item questionnaire screening for obstructive sleep apnea risk, returned a low score in four patients and a medium score in two, indicating low to moderate apnea risk across the cohort. With respect to PTSD etiology, all six participants experienced trauma related to interpersonal violence. Additional trauma types included three patients who experienced accidents, two who experienced earthquakes, and two who experienced sexual assault. Four patients (66.7%) were taking at least one psychotropic medication (defined here as antidepressant, anxiolytic, hypnotic, antipsychotic, or mood stabilizer), including two on SSRIs (33.3%). Additionally, three patients were on hypnotics (50.0%) and three on antipsychotics (50.0%). Per-participant medication details are provided in Supplementary Table S1.

**Table 1.** Participant demographics.

|  | n (%) | Mean(SD); Min-Max |
| --- | --- | --- |
| Female | 6 (100.0%) |  |
| **Age** |  | 40.5(8.3); 29-50 |
| 20-29 years | 1 (16.7%) |  |
| 30-39 years | 2 (33.3%) |  |
| 40 years or over | 3 (50.0%) |  |
| **Job** |  |  |
| Employed | 2 (33.3%) |  |
| Employed (with disability) | 1 (16.7%) |  |
| Not employed | 3 (50.0%) |  |
| **Onset age years old** |  | 20.2(5.7); 10-27 |
| 10-19 years | 1 (16.7%) |  |
| 20-29 years | 5 (83.3%) |  |
| **Stop Bang** |  |  |
| Low (0-2) | 4 (66.7%) |  |
| Medium (3-4) | 2 (33.3%) |  |
| High (5 or over) | 0 (0.0%) |  |
| **PTSD etiology** |  |  |
| Accidents | 3 (50.0%) |  |
| Earthquakes | 2 (33.3%) |  |
| Interpersonal violence | 6 (100.0%) |  |
| Sexual assault | 2 (33.3%) |  |
| Others | 2 (33.3%) |  |
| **Comorbid psychiatric diagnoses** |  |  |
| None | 2 (33.3%) |  |
| Eating disorder | 2 (33.3%) |  |
| Dissociative disorder | 1 (16.7%) |  |
| Bipolar disorder (stable) | 1 (16.7%) |  |
| **Concomitant medications** |  |  |
| Antipsychotic drugs | 3 (50.0%) |  |
| Antidepressant drugs | 2 (33.3%) |  |
| Anxiolytics | 2 (33.3%) |  |
| Mood stabilizers | 1 (16.7%) |  |
| Hypnotics | 3 (50.0%) |  |
| Others | 6 (100.0%) |  |
| **Concomitant therapies** |  |  |
| Counseling | 1 (16.7%) |  |
| CBT | 2 (33.3%) |  |

### Primary Outcome/Safety and Adverse Events

The primary objective of the present study, which was to test the feasibility of the SES procedure in humans, was accomplished. As described above (Figure 2), 6 of the 8 participants who entered the sound-development phase with candidate personalized stimuli prepared (75%) proceeded to Visit 4 and all 6 completed the overnight sleep-based sound stimulation protocol successfully (completion rate 100%). There were no significant protocol deviations affecting the Visit 4 overnight SES intervention or the primary feasibility endpoint. In H08, the home sleep EEG recording device had already been provided at an outpatient encounter when the formal written-consent visit was rescheduled approximately one week later than originally planned; during this unintended scheduling gap, one home recording was inadvertently performed six days before written informed consent was obtained. This recording was excluded from all analyses per GCP, reported transparently, and did not contribute to eligibility determination or endpoint analyses (see also Methods §Screening and Stimulus Development). All six participants completed the protocol-specified procedures (full overnight intervention, and all assessments at Visits 4 and 5) without any participant requesting early termination.

Adverse events, evaluated as secondary endpoints, were few and mild, and none were judged by the study team to be attributable to the auditory stimulation during sleep. Psychiatric AEs were monitored using the Common Terminology Criteria for Adverse Events (CTCAE) v5.0 PTSD-relevant symptom excerpt prespecified in the protocol (see Methods §Adverse Event Monitoring). During the waking stimulus development sessions—in which participants were asked to recall traumatic experiences to create personalized sounds—agitation was indicated in three participants (50.0%; all CTCAE grade 1) and hallucinations in two participants (33.3%; both CTCAE grade 1). These reactions are consistent with the distress inherently associated with trauma recall. On the morning following the overnight procedure (Day 22), three CTCAE grade 1 events were recorded across two participants: one participant (16.7%) experienced transient hallucinations, and another participant (16.7%) reported both mild irritability and a mild headache. All three events were judged by the study team to be related to the underlying disease rather than the SES procedure and resolved within the same day; none of these symptoms were present at the subsequent follow-up visit (Visit 5). Outside the CTCAE PTSD-relevant symptom excerpt, one non-psychiatric adverse event was recorded: one participant (16.7%) developed herpes zoster (CTCAE grade 1) with onset five days before the overnight procedure, which resolved with valaciclovir treatment by the day of overnight stimulation; this event was judged by the study team to be unattributable to a specific cause and was not considered related to the SES procedure. All adverse events recorded throughout the study were CTCAE grade 1; no grade ≥2 events were observed. No cases of confusion, delirium, suicidal ideation, or suicide attempts were observed throughout the study. No worsening in depression was observed from pre-intervention to follow-up, as assessed by the Beck Depression Inventory-II (BDI-II), a 21-item self-report measure of depression severity (mean change 0.83 [95% CI −5.72 to 7.38], p = 0.757). Sleep quality also did not worsen, as measured by the Pittsburgh Sleep Quality Index (PSQI), a self-report questionnaire assessing sleep quality (−1.17 [−8.11 to 5.78], p = 0.684). Similarly, no worsening was observed in the Sheehan Disability Scale (SDS), a measure of functional impairment, across the domains of work/schoolwork, social and leisure activities, and family life/home responsibilities (−0.8 [−2.6 to 1.0], p = 0.289; −1.3 [−3.3 to 0.6], p = 0.140; and −0.3 [−2.8 to 2.1], p = 0.741, respectively). Device-related events were recorded in three participants during the overnight stimulation period but did not preclude protocol completion in any case and resulted in no clinically relevant safety events; the events comprised a non-responsive event-tagging button on the sound-generator console, an EEG-recording PC power insufficiency that occurred after sound delivery had been completed and caused partial recording loss, and an inadvertent activation of an earphone’s built-in environmental-sound playback that was promptly resolved by replacing it with a different earphone. Corrective actions are detailed in the Methods.

### Protocol Fidelity and N3 Targeting

While implemented to verify autobiographical relevance, tolerability, and the SUDs trajectory, the pre-sleep waking exposure to each finalized auditory stimulus is also temporally consistent with the retrieval-extinction paradigm, in which a brief retrieval cue facilitates subsequent extinction learning [35,36], although the present design cannot determine whether retrieval-extinction-like mechanisms are engaged in SES.

The proportion of time that sound stimulation took place during N3 was determined by sleep staging performed according to standard American Academy of Sleep Medicine (AASM) rules, with one modification: the boundaries of the 30-second scoring epochs were aligned to the onset of sound stimulation rather than to the conventional clock-time grid. N3 percentages during sound stimulation ranged from 31.2% (H14) to 88.5% (H09), with a mean of 68.2% (see Table 2). The low N3 percentage for H14 may be explained by contamination of sleep spindles into N3 from N2 which hindered sleep staging. However, overall, this indicates that the protocol enabled successful delivery of the majority of sound stimulation during N3, despite substantial variation among participants (range 31.2–88.5%). Each participant’s personalized stimulus consisted of a 30-second auditory sequence.

**Table 2.**
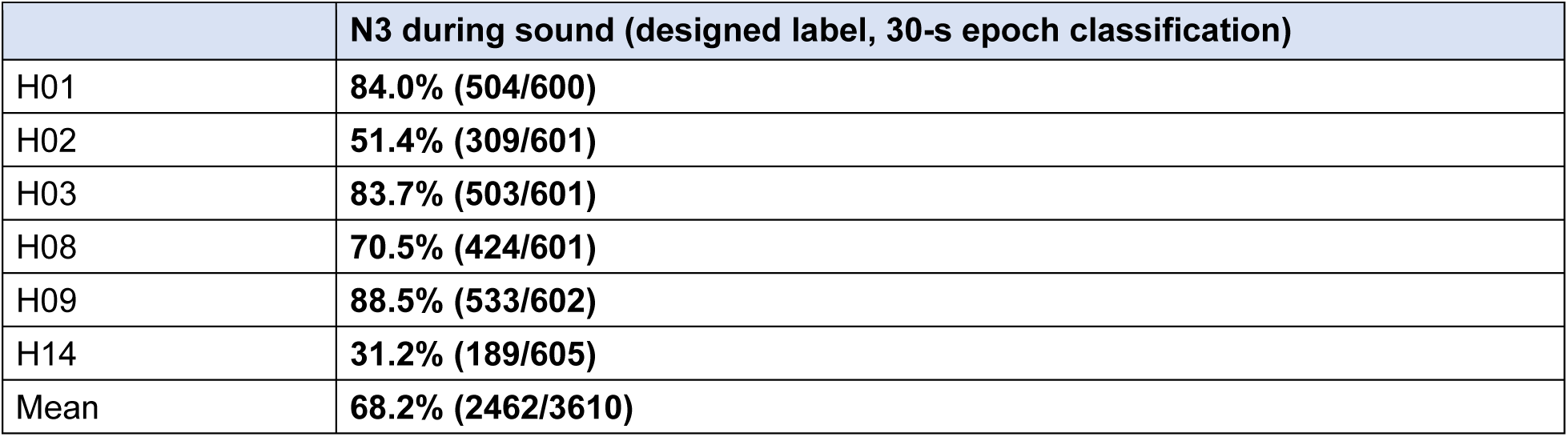
Percentage of sound stimulation during SES that was delivered during N3 sleep. Values in brackets represent the number of seconds of sound stimulation delivered in N3 out of total sound stimulation duration (in seconds).

Immediately prior to sleep, stimulus volume was individually titrated for each participant by presenting a standardized calibration tone through the earphones and adjusting the level downward to the lowest setting at which the tone remained clearly audible. The 30-second sequence was then repeatedly delivered during overnight SES up to a cumulative stimulation duration of 600 seconds (10 minutes).

Importantly, analyses found that the amounts of N3 sleep obtained by participants in the screening phase (in a home environment) and in the P1 unit during sound stimulation did not differ significantly (p = 0.95, paired t-test, two-tailed; Figure 3h). Total sleep time, total time in N1, and total time in N2 were significantly greater in the P1 unit compared with the home screening night (TST: p = 0.045, Figure 3a; N1: p = 0.028, Figure 3f; N2: p = 0.007, Figure 3g; paired t-test, two-tailed), consistent with extended overnight monitoring in a controlled clinical setting. Sleep latency, sleep efficiency, arousal index, wake after sleep onset, REM duration, and REM latency did not differ significantly between conditions (sleep latency: p = 0.085; sleep efficiency: p = 0.207; arousal index: p = 0.232; WASO: p = 0.147; REM duration: p = 0.675; REM latency: p = 0.135; Figure 3b–e, i, j). Given this result, the present results support the feasibility of N3-targeted delivery under monitored conditions.

**Figure 3.**
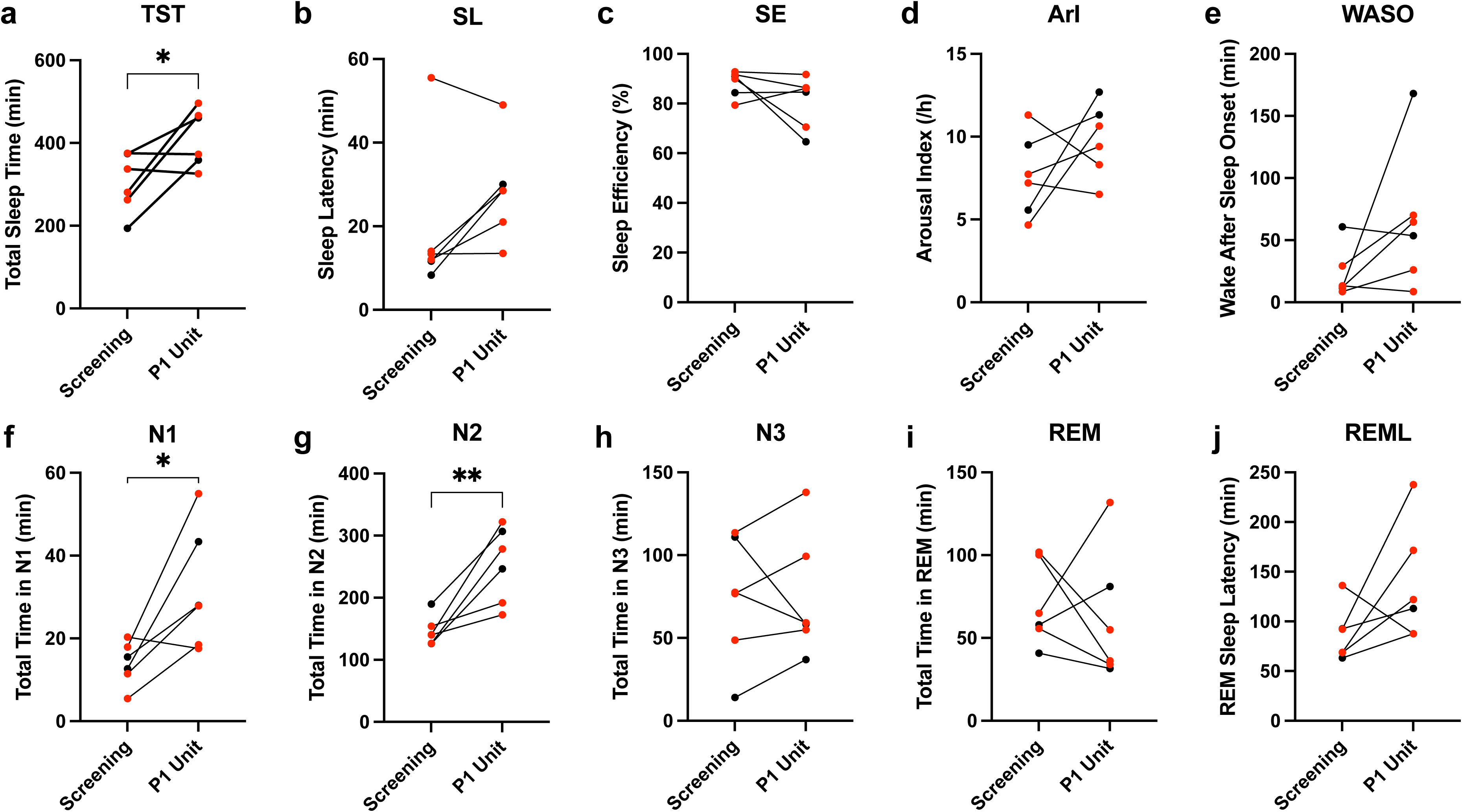
Sleep architecture metrics comparing screening at home to the SES procedure in the P1 unit. Individual participant data are shown for (a) total sleep time, (b) sleep latency, (c) sleep efficiency, (d) arousal index, (e) wake after sleep onset, (f) total time in N1, (g) total time in N2, (h) total time in N3, (i) total time in REM sleep, and (j) REM sleep latency, measured in minutes during the home screening night and the night in the P1 unit during the SES procedure. Each line represents an individual participant. Black dots indicate participants who received Version A of the protocol; red dots indicate participants who received Version B of the protocol. Significant differences between conditions are indicated by brackets: *p < 0.05, **p < 0.01 (paired t-test, two-tailed). Screening values are averaged across post-consent home-recording nights (H01, H02, H03, H08: 3 nights; H09: 4 nights; H14: 3 nights). For H08, the 2025-03-26 home recording, which preceded written informed consent (2025-04-01), was excluded from all screening-phase averages; the screening average for H08 is therefore based on 3 post-consent nights (2025-04-04, 04-05, 04-06).

### SUDs

Participants demonstrated differential SUDs change in relation to the protocol version assignment (Figure 4). Participants who received the Version B protocol (H03, H08, H09, H14), in which the sound selected for overnight sleep delivery was permitted to elicit any SUDs level (no ceiling imposed), showed reductions in subjective distress (SUDs ratings) from pre- to post-SES that reached nominal statistical significance in this exploratory analysis (p < 0.05). These participants showed a mean SUDs reduction of 50.0% (i.e., mean SUDs change = −50.0%; standard deviation [SD], 24.5%). In contrast, those who received the Version A protocol (H01, H02) showed a trend toward slight worsening in SUDs (mean SUDs change = +15.5%; SD = 1.7%, under the unified sign convention in which positive values indicate post-procedure worsening). Under Version A, the sound delivered during sleep was selected from candidates eliciting SUDs of 30–40, although candidates outside this range were inevitably encountered earlier in the sound-development process. The difference between the two groups was estimated at −65.48% (95% confidence interval, −104.24% to −26.72%; nominal p = 0.012, Welch t-test). Version B participants showed a numerically larger exploratory decrease in SUDs compared with Version A; sham-controlled confirmation is required before inferring a treatment effect.

**Figure 4.**
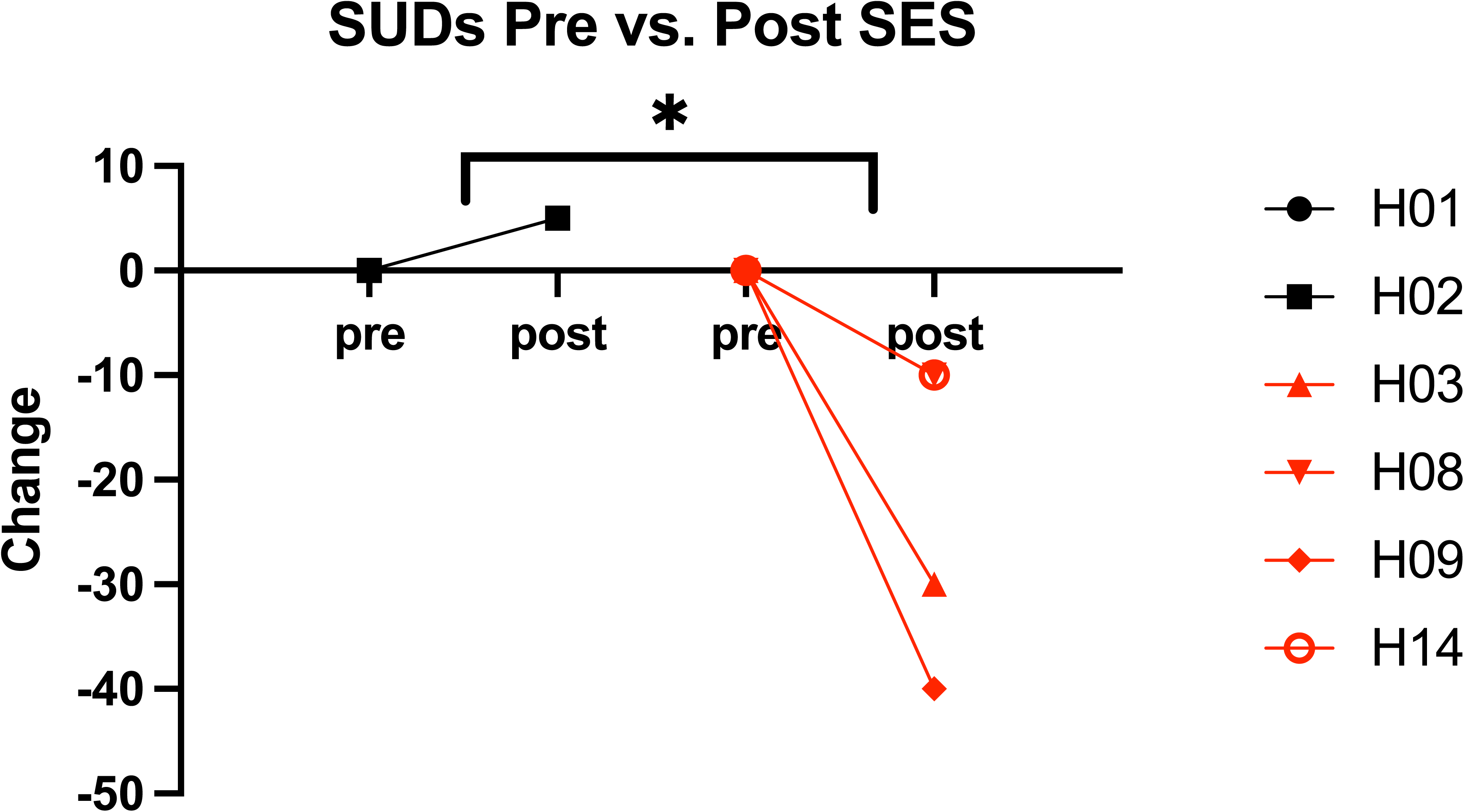
Pre-post changes in SUDs following SES. Individual participant data are shown for SUDs ratings obtained during sound exposure before and after the SES procedure. Participants receiving Version A are denoted in black and Version B are denoted in red. Data are expressed as change from baseline. Pre-SES values were normalized to 0, and post-treatment values represent the absolute SUDs-point difference between post- and pre-treatment (post − pre). The asterisk reflects between-version comparison performed on the proportional change ((post − pre) / pre × 100%) as defined in Methods §Statistical Analysis; raw point values are shown here to allow inspection of per-participant magnitudes. Significant differences are indicated by brackets: *p < 0.05 (Welch t-test, two-tailed; nominal exploratory).

### Physiological Measures

To probe whether sleep-time delivery of trauma-linked auditory cues engages autonomic circuits classically activated by waking trauma-cue exposure in PTSD, we examined heart rate variability (HRV) and skin conductance level (SCL) during the sound stimulation period. During wakefulness, exposure to personalized trauma cues reliably elicits a well-documented autonomic signature in individuals with PTSD, characterized by increased SCL and decreased HRV, reflecting a strong sympathetic shift and parasympathetic withdrawal [37–39]. However, autonomic responsiveness is profoundly altered by sleep states; specifically, slow-wave sleep (SWS) imposes robust parasympathetic dominance [40], and targeted memory reactivation (TMR) paradigms typically deliver sensory cues at low intensity over a white-noise background to remain below an arousal threshold [34]; under such stably-asleep conditions, substantial autonomic perturbations would not be expected. We a priori hypothesized that if Sound Exposure during Sleep (SES) successfully mobilizes the trauma memory representation during SWS, SCL and HRV might exhibit partial signatures of this waking response; alternatively, if the sleep state fully attenuates this reactivity, autonomic measures may remain at the sleep-state baseline. We therefore analyzed HRV (RMSSD, SDNN) and SCL across the entire 600-second stimulation period (the full duration of overnight sound delivery).

Two HRV parameters (RMSSD and SDNN) collected during the sound stimulation period were analyzed across three 200-second time windows (Figure S1). Pearson correlations between SUDs change and HRV were not statistically significant for either parameter in any window (SDNN: First 200s r = +0.46, p = 0.36; Second 200s r = −0.14, p = 0.80; Third 200s r = −0.10, p = 0.85. RMSSD: First 200s r = +0.46, p = 0.37; Second 200s r = −0.15, p = 0.78; Third 200s r = −0.11, p = 0.84).

We examined the correlation between SUDs change scores [(Post − Pre) / Pre] and average standardized skin conductance level (SCL) across three 200-second time windows during the 600-second stimulation period (n = 5; H03 was excluded due to SCL recording failure; Figure S2).

A trend emerged in which greater improvement in SUDs (i.e., more negative SUDs change scores under the unified sign convention) was descriptively associated with higher standardized SCL, with the relationship differing across time windows. Pearson correlations indicated the strongest (negative) association during the middle 200-second window (Second 200s: r = −0.80, p = 0.104), where participants whose SUDs change scores were more negative (approaching −1.0) also demonstrated elevated standardized SCL. The first and third 200-second windows showed weaker and inconsistent relationships (First 200s: r = −0.60, p = 0.29; Third 200s: r = +0.28, p = 0.65). Participant H02 showed notably elevated standardized SCL despite modest SUDs change, while H01 and H09 clustered more closely along the middle window regression line. However, these correlations did not reach statistical significance (p>0.05).

As an exploratory analysis, we additionally examined the relationship between SUDs change and the percentage of time spent in N3 sleep during the SES procedure. Visual inspection across Version A, Version B, and the combined sample did not reveal a consistent relationship; patterns differed between protocol versions and no statistically significant correlation was observed (Figure S3).

### PCL-5

PCL-5 scores (self-report measure of PTSD symptom severity over the preceding week) were collected at all study visits (Visits 0 through 5). The present analysis compared the pre-SES (Visit 4, immediately before sleep stimulation) and follow-up (Visit 5) scores, in accordance with the recommended ≥1-week PCL-5 administration interval. Analysis of PCL-5 sum scores showed a trend toward decreases in participants who received Version B, whereas those receiving Version A showed minimal change (Figure 5a). The mean change in total PCL-5 scores was −13.5 (SD = 10.4) versus 0.5 (SD = 0.7), with an estimated mean difference of −14.0 (95% CI, −30.4 to 2.4; nominal p = 0.073, Welch t-test). The Intrusion subcategory score revealed a reduction in Version B participants from pre- to post-SES that reached nominal statistical significance in this exploratory analysis, with a mean change of −5.0 (SD = 2.4; nominal p = 0.015, Welch t-test; Figure 5b). In contrast, participants receiving Version A showed a mean of 2.0 (SD= 1.4). The between-version difference in Intrusion was −7.0 (95% CI, −11.6 to −2.4; p = 0.015); consistent with the feasibility framing, this is reported as a hypothesis-generating exploratory signal rather than confirmatory evidence of efficacy. The Avoidance subcategory showed similar trends toward reduction in Version B, although not statistically significant (Figure 5c). No statistical significance was found in analyses of the Negative Alterations (Figure 5d) and Hyperarousal (Figure 5e) categories (p>0.05).

**Figure 5.**
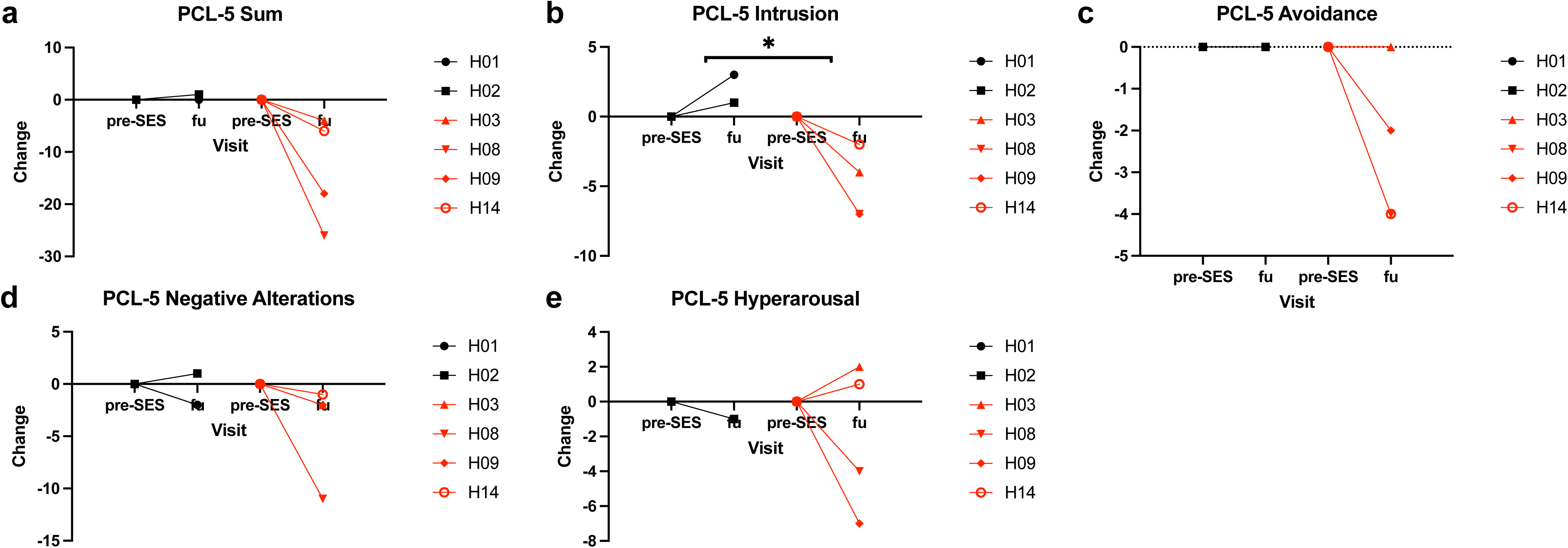
Pre-SES to follow-up (fu) changes in PCL-5 scores. Individual participant data are shown for (a) sum, (b) intrusion, (c) avoidance, (d) negative alterations, and (e) hyperarousal PCL-5 scores obtained at Visit 4 (prior to the overnight SES procedure) and at follow-up (Visit 5). Participants receiving Version A are denoted in black and Version B are denoted in red. Data are expressed as change from baseline. Pre-SES scores were normalized to 0, and follow-up scores represent the raw point difference between follow-up and pre-SES PCL-5 scores (fu − pre). Significant differences are indicated by brackets: *p < 0.05 (Welch t-test, two-tailed; nominal exploratory).

In addition to PCL-5, we administered the Posttraumatic Diagnostic Scale for DSM-5 (PDS-5), which captures symptom changes over a one-month window. Overall trajectories were broadly consistent with PCL-5—Version B participants showed greater symptom reductions than Version A—but the between-version difference did not reach statistical significance (mean difference −8.5 points; 95% CI −22.3 to 5.3; Figure S4a-e). Because the present study evaluated the acute effects of a single-night intervention, PCL-5 (past-week) and PDS-5 (past-month) capture different temporal dimensions of PTSD symptom change and are not directly interchangeable; the former captures short-latency change, while the latter integrates pre-intervention baseline days. Pre-intervention symptom stability was also assessed: SUDs measured at Visit 2 versus the pre-SES timepoint (Visit 4 Day 21) did not differ significantly (n = 6; paired t = 1.00, p = 0.363), and PCL-5 intrusion scores across Visit 0 (home screening), Visit 1, and Visit 2 likewise showed no significant change (n = 6; Friedman χ² = 3.27, p = 0.195; pairwise paired t-tests all p > 0.05; Figure S5), indicating no statistically detectable pre-intervention change, although the study was underpowered to exclude spontaneous fluctuation.

### Sensitivity Analyses for Statistical Robustness

Because version assignment and enrollment date are near-perfectly collinear (Pearson r = 0.96), we conducted sensitivity analyses that assess statistical robustness without disentangling version from calendar time (the temporal-confounding limitation is addressed in the Limitations section of the Discussion; sequential version implementation shown in Figure S6). The observed between-version difference was the single most extreme mean difference attainable in the exact permutation distribution (15 possible splits); the corresponding exact two-sided p-value was 0.067 for both PCL-5 intrusion and SUDs, which does not reach α = 0.05 due to the combinatorial floor of 1/15 in this design. The nominal Welch p-values (0.012, 0.015) should therefore be interpreted as descriptive only. Leave-one-out analysis for PCL-5 intrusion change retained p < 0.05 in all four evaluable drops (range 0.015–0.037; LOO was restricted to Version B drops because Version A contained only n = 2 participants, which would leave n_A = 1 and render the Welch standard error undefined), with the effect estimate remaining within ±15% of the full-sample estimate; for SUDs change, two of four LOO drops crossed the conventional α = 0.05 threshold (p range 0.016–0.063)—that is, removing certain individuals shifted the result above or below the 5% significance boundary, indicating that the SUDs effect estimate is fragile to single-participant exclusion. Spearman correlations between each outcome and enrollment month were all negative under the unified sign convention (negative values indicate improvement), consistent with later-enrolled participants showing larger improvements (PCL-5 intrusion r = −0.65, p = 0.16; PCL-5 total r = −0.75, p = 0.08; SUDs r = −0.60, p = 0.21) but did not reach conventional significance with n = 6. Within-Version-B Spearman correlations (n = 4) are reported descriptively only in Supplementary Table S2. Notably, response was observed across heterogeneous trauma types—including the participant with earthquake-related trauma (H08), who showed a clear PCL-5 intrusion reduction—suggesting that the SES effect may not be confined to a single trauma category, though extrapolating generalizability from this single-case observation is strictly anecdotal.

## Discussion

In this study, we demonstrate the feasibility of trauma-associated auditory stimulation during SWS and provide preliminary human evidence that delivering trauma-linked cues during SWS is associated with reductions in core PTSD intrusion symptoms. Building on prior TMR paradigms that have employed cues associated with experimentally conditioned stimuli or therapy-linked signals (e.g., EMDR-paired clicks) [28], our approach delivered cues intrinsically tied to each participant’s autobiographical trauma during SWS. While the present feasibility study cannot directly establish that the trauma memory ensemble itself was reactivated, these findings position autobiographical-trauma-cued SWS stimulation as a candidate framework for the future development of stand-alone, sleep-based PTSD interventions.

The primary aim of this study was to establish whether SES could be safely and practically implemented, and several findings support its feasibility. All six participants who entered the sleep-based sound stimulation phase completed the protocol. Adverse events were few and mild: transient agitation and hallucinations occurred only during the waking stimulus-development sessions—reactions consistent with trauma recall itself—while the overnight SES procedure produced no adverse events judged by the study team to be attributable to sleep-based stimulation.

A central concern surrounding sleep-based memory reactivation is the potential for disruption of sleep architecture or exacerbation of distress, given that SWS is itself critical for memory consolidation. However, the sleep architecture data support the safety of the intervention. Total sleep time increased during the SES night relative to baseline home sleep, and critically, SWS duration—the stage targeted for cue delivery—did not differ from baseline, indicating that auditory stimulation did not suppress SWS. Although increases in N1 and N2 durations were observed, with a non-significant trend in sleep latency, these stages were not targeted for sound delivery and the changes did not appear to compromise SWS stability.

As reported in Results (see Figure S5), both SUDs and PCL-5 intrusion scores remained stable across pre-intervention visits, with no spontaneous improvement detected. This stability is notable given the chronicity of symptoms in the cohort and may suggest that the observed post-SES reductions do not solely reflect natural symptom variability or regression to the mean, although the study was underpowered to exclude these alternative explanations. We therefore present the post-SES reductions as a hypothesis-generating signal, while sham-controlled studies are required to distinguish sleep-based memory-reactivation effects from expectancy, regression to the mean, and other nonspecific factors.

These feasibility findings have direct implications for scalability. Recent advances in consumer-grade wearable EEG devices with automated sleep staging algorithms—including closed-loop TMR implementations feasible outside research sleep laboratories [30]—together with auditory VR platforms for rapid stimulus generation [41], may enable patients to receive SES at home without overnight hospitalization, substantially reducing cost and logistical barriers that currently limit access to evidence-based PTSD treatments [3]. However, home-based implementation will require the development of sound stimuli optimized for sleep-based delivery, as well as remote safety monitoring protocols. Furthermore, the device malfunctions reported above (3 of 6 participants) underscore the importance of validating hardware reliability in larger monitored samples.

Our framework identifies and deploys auditory stimuli intrinsically linked to the individual’s traumatic experience through structured clinical interview, with the intent of engaging the trauma-related memory representation during SWS without requiring an intermediate step of linking neutral cues (e.g., clicks or tones) to trauma memories through conditioning or therapeutic procedures, as employed in prior TMR studies of PTSD and related conditions [28]. Notably, prior TMR in PTSD demonstrated that auditory stimulation during SWS is technically feasible and safe in this population [28]. That study employed EMDR-paired clicks as reactivation cues and compared the intervention to a silent control condition. In the present study, the use of content-specific, autobiographical trauma-linked auditory stimuli—rather than therapy-procedure cues paired with an active-sound-versus-silent contrast—is intended to inform the question of whether the memory content carried by the cue, not auditory stimulation per se, may be associated with extinction-like mechanisms during SWS. Careful stimulus selection and validation are central to this approach, as cue salience and specificity likely determine the degree of trauma-memory reactivation achieved during SWS. Scalable implementation will require efficient methods for generating and standardizing trauma-relevant auditory cues without compromising specificity. Recent work by Yamauchi et al. addresses this scalability challenge by developing an LLM-driven, natural-language interface that enables clinicians to generate patient-specific auditory VR stimuli within a single clinical session, pointing toward a practical pipeline for individualized SES cue production [41].

Beyond technological scalability, another question concerns the degree of personalization required to reliably engage trauma-related networks. Although personalized cues preserve autobiographical specificity, converging evidence suggests that category-consistent (“generalized”) trauma cues can similarly activate disorder-relevant circuits. Virtual reality exposure systems such as Virtual Iraq/Virtual Afghanistan employ standardized libraries of combat-related sights and sounds rather than individualized autobiographical stimuli [42]. Despite this standardization, substantial symptom reductions and sustained improvements have been reported [43]. These findings indicate that carefully designed, category-matched cues can support clinically meaningful extinction even without full autobiographical tailoring.

Neuroimaging research further supports this view. Gramlich and colleagues demonstrated that a standardized trauma-related sound (an explosion) specifically activated the right medial superior prefrontal cortex—a region central to threat regulation—in veterans with PTSD, and that this response was absent for non-trauma sounds [44,45]. Additional support comes from the International Affective Digitized Sounds (IADS) database, which demonstrates highly reliable valence–arousal ratings across large and cross-cultural samples [46–50]. Unpleasant high-arousal sounds consistently cluster within the negative-valence quadrant [51], underscoring the stability of affective responses to standardized stimuli. Similar disorder-specific hyperreactivity to non-personalized auditory triggers is observed in misophonia and specific phobias [52–56]. Collectively, these findings suggest that sufficiently salient category-consistent cues can recruit pathological memory or threat-processing networks even without full autobiographical customization.

Building on this body of work, our stimulus-selection protocol was deliberately designed to avoid sounds that, on independent normative grounds, would be expected to elicit distress or arousal in healthy listeners—for example, abrupt loudness changes or piercing/screech-like spectral profiles—even when such sounds might appear thematically related to a given trauma. Existing normative databases such as IADS provide validated valence–arousal coordinates for a wide range of standardized sounds and thereby make it operationally feasible to systematically exclude such candidates during SES sound development, supporting an approach in which content-specific stimulation can be pursued without inadvertently introducing nonspecific aversive arousal.

SES is grounded in emotional processing theory (EPT), which holds that therapeutic change requires activation of the pathological fear structure and integration of safety information that competes with the original threat associations [22,57,58]. Consistent with this framework, rodent engram research has identified distinct neuronal ensembles for fear and extinction memories that compete for behavioral expression [59,60]. The post-procedure changes observed here may be compatible with a sequence in which the sound-development phase provided an initial, incidental safety contextualization within a secure clinical environment, and re-presentation of these cues during SWS—a state of heightened memory plasticity—may have provided conditions compatible with updating of the safety-associated trace [27,61,62]. The present uncontrolled design cannot establish this mechanism.

Notably, TMR has also shown therapeutic promise beyond SWS-based interventions: Schwartz and Perogamvros demonstrated that playing a sound previously associated with a positive dream scenario during REM sleep, in combination with imagery rehearsal therapy, significantly reduced nightmare frequency and distress in patients with nightmare disorder [29]. This finding underscores the versatility of sleep-stage-targeted auditory reactivation across different clinical applications.

The temporal sequence of SES—a brief waking exposure to the personalized trauma-linked sound followed several hours later by repeated presentations of the same stimulus during SWS—shows surface temporal resemblance to the retrieval-extinction paradigm [35,36]. In that classical framework, a brief reminder cue reopens a post-retrieval reconsolidation window, rendering the memory labile and susceptible to update via subsequent unreinforced exposure. While SES substitutes traditional awake extinction with sleep-state targeted memory reactivation, the temporal architecture of the protocol aligns with this core sequence of a discrete waking retrieval followed by non-threatening stimulus re-exposure. We note critical caveats: the interval between waking retrieval and overnight SWS exposure approaches the canonical boundary of the reconsolidation window (∼6 hours) [36], SWS presents a fundamentally different neuromodulatory environment than waking extinction, and our trial lacks the component controls necessary to isolate reconsolidation effects. This cross-state parallel is therefore offered as a speculative analogy only, not as a replacement for the EPT-based safety-signal framework that grounds our primary interpretation; component-controlled testing would be required before adopting it as a mechanistic explanation.

### Placebo and Nonspecific Therapeutic Factors

A central interpretive limitation of this open-label, single-arm design is the absence of a sham comparator. Each participant underwent two bi-weekly clinic visits devoted to the collaborative development of a personalized trauma-associated sound, during which repeated therapeutic contact, instillation of hope, and positive expectancy were unavoidably present. In randomized trials of established PTSD psychotherapies, nonspecific therapeutic factors—including clinician attention, treatment credibility, and patient expectancy—have been estimated to account for a substantial proportion of observed symptom change. Consequently, the present findings cannot adjudicate between a sleep-specific memory-reactivation mechanism and a nonspecific placebo-expectancy mechanism. Future confirmatory trials should incorporate an active sham arm—for example, auditory stimulation matched for loudness, frequency content, and clinical contact but using scrambled or affectively neutral sounds—together with blinded assessment of clinical outcomes, to isolate the contribution of SWS-timed autobiographical trauma reactivation.

### Content-Specificity of Auditory Stimulation

A related limitation is that our design cannot isolate the contribution of trauma-specific auditory content from that of auditory stimulation per se. Given that generic auditory stimulation during SWS is known to alter sleep architecture—such as modulating slow-oscillation activity and slow-oscillation-spindle coupling—we cannot exclude that the observed reductions in intrusion symptoms reflect nonspecific neurophysiological effects of any sound presented during SWS, rather than properties intrinsic to personalized trauma-associated cues. This nonspecific-auditory contribution can be dissociated from content-specific stimulation within the same active-sham randomized design proposed under Placebo and Nonspecific Therapeutic Factors.

Several limitations should be considered. First, device malfunctions involving the sound-generator console, EEG-recording PC, and earphones were reported in three participants. Second, the small sample size (n = 6) reflects the feasibility-focused design of this open-label pilot study and limits statistical power. This restricted cohort size also limits trauma-type representation and reflects selection bias inherent in physician-judged eligibility, requiring broader inclusion criteria in future studies. The unequal and non-randomized distribution between protocol versions (Version A, n = 2; Version B, n = 4) further constrains statistical inference. As noted above, the comparison between protocol versions is subject to temporal confounding arising from their sequential implementation: Version A participants (n = 2) were enrolled in September–October 2024, whereas Version B participants (n = 4) were enrolled between March and June 2025, leaving a four-month gap between the last Version A and the first Version B enrollment (Supplementary Fig. S6). Potential confounders that co-vary with this timeline include seasonal factors, accumulating operator experience, evolving selection patterns, and the mid-trial protocol amendment itself. Version assignment and enrollment month are therefore near-perfectly collinear (Pearson r = 0.96), precluding meaningful decomposition of version and time effects by analysis of covariance. Sensitivity analyses (Supplementary Table S2) descriptively support the observed separation but cannot distinguish protocol-version effects from temporal mechanisms; the present effects should be interpreted as preliminary. The all-female composition of the cohort (100% female; n = 6) further limits generalizability across sex; future controlled trials should include both sexes to assess potential sex-related differences in feasibility and tolerability.

Third, half of the cohort (50%; n = 3 of 6: H01, H08, H14) were on stable memantine therapy at enrollment, with doses and regimens unchanged throughout the trial period. Memantine is an N-methyl-D-aspartate receptor antagonist, a mechanism directly implicated in the synaptic plasticity, memory reconsolidation, and fear-extinction processes that SES is designed to engage, and a small open-label clinical study has explored its use as adjunctive PTSD pharmacotherapy [63]. Although the present design did not permit isolation of any memantine contribution, this background pharmacotherapy represents a substantive mechanistic confounder for any SES-related symptom change observed. We further note that the CRF captured in-trial continuity rather than precise pre-enrollment medication start dates, meaning the exact duration of stable memantine exposure prior to Visit 0 cannot be quantified; however, the aforementioned lack of any in-trial dose adjustments confirms that no active dose titration occurred during the SES protocol. A descriptive sensitivity analysis comparing exploratory symptom outcomes and pre-treatment baselines by memantine status (n = 3 on memantine vs. n = 3 not on memantine) is provided in Supplementary Table S3; given the very small subgroup sizes, formal hypothesis testing is mathematically underpowered (minimum two-sided exact permutation p = 0.10) and is not reported. Future controlled trials should pre-specify NMDA-modulating medication strata or exclude such concomitant medications to disentangle SES-specific from pharmacological contributions.

Finally, the autonomic readouts reported above must be interpreted within the physiological constraints of sleep-state recording. The lack of significant HRV changes and the only non-significant descriptive association between more negative SUDs change scores and higher standardized SCL likely reflect SWS-dominant parasympathetic tone, which naturally attenuates the autonomic cascades that manifest during wakefulness [40]. Furthermore, extracting brief-window RMSSD from wrist-worn photoplethysmography is subject to well-documented precision limitations relative to ECG, which restricts the statistical power to detect subtle micro-autonomic shifts [64,65]. From a translational perspective, achieving distress reduction (SUDs) via targeted memory reactivation without precipitating the overwhelming somatic burden typically associated with waking exposure therapies is consistent with a favorable safety and tolerability profile for SES.

Future Directions and Required Confirmatory Design. Resolving the open questions identified above will require an integrated confirmatory trial design that combines (i) randomized assignment to an active sham-sound arm matched for loudness, spectral content, and clinical contact; (ii) randomized assignment to higher- versus lower-SUDs stimulus-intensity conditions; and (iii) blinded outcome assessment to minimize detection bias, particularly for subjective measures such as SUDs. Such a single, integrated design simultaneously dissociates content-specific stimulation from nonspecific auditory effects, separates a sleep-specific memory-reactivation mechanism from nonspecific placebo and expectancy effects, and resolves the version × time confound arising from sequential implementation of the two protocol versions.

Taken together, although preliminary, these findings establish feasibility and provide initial support for the hypothesis that trauma-linked auditory reactivation during SWS may be compatible with mechanisms relevant to pathological fear processing, warranting systematic evaluation in larger controlled trials.

## Methods

### Participants

The sample size was determined based on feasibility considerations. With six participants, the study had an 88% probability of detecting at least one adverse event if the true incidence was 30%. To account for the possibility that some participants could not make the stimulus sound, the target sample size was set to 6∼12 participants. This calculation pertains solely to feasibility and safety endpoints; the study is not powered to detect efficacy-related differences. Accordingly, all between-group comparisons of symptom-related outcomes should be interpreted as exploratory and hypothesis-generating, intended to inform effect-size estimation for a subsequent adequately powered randomized controlled trial. Thirteen adult patients were enrolled through referrals from multiple psychiatric care providers in Japan, including the National Center of Neurology and Psychiatry (NCNP) and Hori Mental Clinic. Inclusion criteria included regular psychiatric care under a confirmed PTSD diagnosis (with eligibility threshold defined as a Posttraumatic Diagnostic Scale for DSM-5 [PDS-5] total score ≥ 28 at screening, corresponding to moderate-to-severe symptom severity per Foa et al., 2016), sufficient bilateral hearing, the capacity to recall traumatic memories via auditory cues, and ≥ 10 minutes of nightly SWS confirmed through home EEG. Exclusion criteria comprised severe suicidal ideation, psychosis, uncontrolled bipolar disorder, epilepsy, severe physical illness, untreated sleep disorders, uncontrolled hypertension, or insufficient Japanese language proficiency. Ethical approval was obtained from the NCNP Ethics Committee (approval no. B2023-118), and all procedures adhered to the Declaration of Helsinki. Written informed consent was obtained from all participants. The trial was prospectively registered at the Japan Registry of Clinical Trials (jRCT1030230706; registration date 2024-03-15) prior to first participant enrollment, in accordance with ICMJE recommendations.

### Screening and Stimulus Development (Visits 0–3)

At Visit 0, performed at home, eligibility was confirmed using overnight recording with a portable EEG system (see “Equipment and Data Handling”), requiring at least 10 minutes of nightly SWS. For one participant (H08), an additional first overnight recording (2025-03-26) was inadvertently obtained six days before written informed consent (2025-04-01) and was excluded from all analyses; the H08 screening-phase sleep architecture values reported here and in Figure 3 are averages of the three post-consent home-recording nights (2025-04-04, 04-05, 04-06). For participants who met this criterion, baseline clinical assessments (PCL-5, BDI-II, PSQI, SDS, and the STOP-BANG questionnaire) and a trauma-recall questionnaire were also obtained at Visit 0. Participants then attended up to three bi-weekly clinic sessions (Visits 1–3) to develop and refine trauma-recalling sounds based on anxiety-provoking scenarios. Sessions were held in private clinical or secure rented rooms. Candidate sounds (≤30 seconds, ≤50 dB) were evaluated using the Sound Trauma Similarity Questionnaire, and clinicians ensured each sound’s safety and tolerability. Two versions of the protocol were administered. This study was originally designed as the first human trial of auditory stimulation during sleep for PTSD patients. Therefore, for safety reasons, the sound ultimately selected for overnight sleep delivery in the first two participants was constrained to SUDs of 30–40, even though candidate sounds outside this range were necessarily encountered during the development phase prior to final selection. After completion of the first two cases, however, no adverse events were judged by the study team to be attributable to the auditory stimulation, including no arousal responses in EEG during sound presentation. Following completion of overnight stimulation in the first two participants (H01 V4 = 2024-10-26; H02 V4 = 2024-12-21) with no adverse events judged by the study team to be attributable to the sleep-based auditory intervention (and no arousal responses in EEG during sound presentation in either participant), a formal protocol amendment was prepared and submitted to the NCNP Ethics Committee (Form 3 protocol-comparison record v1.91; received by the IRB office on 2025-01-27 as part of the amendment package subsequently registered under NCNP receipt 2024-1428). The amendment replaced the prescribed SUDs 30–40 sound-selection rule with physician multi-criteria sound determination governed by the IRB-approved Sound Stimulation Go/NoGo Criteria attachment v2.0—a structured clinical assessment instrument that integrates Sound Trauma Similarity Questionnaire scores with clinician-observed stress responses. The instrument itself is held under sponsor confidentiality pending an international patent application (domestic filing 特願 2026-027927, 2026-02-24); structural details of the instrument are therefore not disclosed publicly, but its application during this study was conducted exclusively in accordance with the IRB-approved attachment, and full details are available to qualified researchers and editorial reviewers from the corresponding author upon reasonable request under appropriate non-disclosure terms. The amendment was IRB-approved on 2025-02-17 (NCNP receipt 2024-1428; protocol version 2.1; study implementation permit issued 2025-02-20), then rolled forward into protocol version 2.2 (NCNP receipt 2025-033, IRB-approved 2025-03-04, study implementation permit 2025-03-10), under which Version B procedures were conducted between March and August 2025. The two versions are referred to herein as Version A and Version B. H03 (the first Version B participant) provided written informed consent on 2025-03-05 under protocol version 2.1 (study implementation permit issued 2025-02-20), whereas all subsequent Version B procedures—including H03 Visit 4 on 2025-04-19—were conducted after protocol version 2.2 implementation on 2025-03-10. The remaining Version B participants (H08, H09, H14) provided consent and underwent all procedures entirely under protocol version 2.2. Version A (H01, H02) was conducted entirely under the pre-amendment protocol versions, in which the sound ultimately selected for overnight sleep delivery was constrained to elicit SUDs of 30–40. Although candidate sounds outside this range were inevitably generated and tested earlier in the sound-development process, the final sound selection adhered to the SUDs 30–40 rule for both participants. Version B (H03, H08, H09, H14) was conducted entirely under the post-amendment protocol versions and applied the IRB-approved physician multi-criteria sound-determination instrument without any fixed SUDs ceiling on the sound selected for sleep delivery. No other aspects of the protocol varied between Version A and Version B. Sounds were interspersed with neutral, non-traumatic stimuli, and all participants made final selections by Visit 2; Visit 3 was therefore not required.

Clinical assessments across the study used the PCL-5 [66], PDS-5 [67], BDI-II [68], PSQI [69], SDS [70], and SUDs [71]; administration timing is specified for each visit (see also Fig. S4). All concomitant medications were documented on the enrollment Concomitant Medication Case Report Form (CRF) at Visit 0 as “in use prior to study enrollment” (i.e., already being taken at the time of trial registration); continuation status was reverified at every subsequent scheduled study visit (Visits 1–5) via structured concomitant-medication review on the same CRF, and no medication was initiated, discontinued, or dose-adjusted during the SES protocol or the follow-up period (see Supplementary Table S1).

### Sound Selection

The general flow of sound selection for all participants is depicted in Figure 1. For each enrolled participant, personalized auditory stimuli reminiscent of contexts associated with the individual’s reported trauma history were developed in collaboration with the study clinicians across the bi-weekly sound-development sessions (Visits 1–3). At Visits 1 and 2, candidate sounds were presented to the participant and rated for subjective distress (SUDs) via a structured questionnaire, after which the attending clinician issued a Go/No-Go judgment determining whether each candidate would proceed to a further iteration of sound development or to the overnight stimulation session. Specific per-participant sound descriptions are not disclosed publicly to minimize re-traumatization risk in trauma-affected readers; details are available to qualified researchers upon reasonable request, subject to ethics committee approval. All finalized auditory stimuli were prepared as 30-second sound sequences and were identical in duration across all six participants and across both protocol versions.

H04 was not enrolled due to a PDS-5 score that was below the cut-off for study eligibility. H05 had a trauma case of DV but withdrew during the sound-development phase before any SES stimulation sound could be finalized for overnight delivery. H06 had earthquake trauma and entered the sound-development phase with candidate sounds prepared, but was determined to be ineligible at the confirmatory recording due to insufficient SWS, before a final SES stimulation sound was reached. H07 was an unused ID due to a technical issue. Finally, cases H10–H13 were deemed ineligible for the study based on various exclusion criteria.

### Sleep Stimulation (Visit 4)

Participants spent one night at the Phase 1 Unit, Clinical Research Promotion Center, The University of Tokyo Hospital. A sleep diary was completed during the week preceding the overnight session. Prior to sleep, participants re-listened to their 30-second personalized trauma-associated auditory sequence to confirm tolerance and provide pre-SES SUDs ratings; the PCL-5, PDS-5, BDI-II, PSQI, and SDS were also reassessed. Because hearing thresholds vary between individuals, the maximum stimulation volume was individually calibrated immediately before sleep onset: with the participant wearing silicone earphones, a common calibration tone was presented and the minimum volume at which the participant clearly perceived the tone was set as that participant’s volume ceiling, defined as “100%” on the system volume scale throughout the present study. Participants were then fitted with EEG electrodes, skin conductance sensors, and pulse monitors. Auditory stimulation was operated manually by the on-site sleep technician under the audio-playback specification fixed on 11 September 2024, which governed all six SES sessions in this study (Visit 4 dates: 26 October 2024 – 17 August 2025); the on-site technician monitored sleep stage continuously and issued every decision to start, pause, or stop stimulation, while a parallel AI sleep classifier developed for technical validation did not control sound delivery. Thirty-second epochs were defined consecutively from the moment slow-wave sleep was first identified, and an epoch was classified as “N3” when ≥90% of its 30 seconds were judged as N3 by the technician (entered via the SPACE key). When three consecutive N3 epochs accumulated (90 seconds of sustained SWS), playback of the participant-specific 30-second trauma-associated auditory file began at the start of the next epoch, with volume rising from 20% of the calibrated participant-specific scale by +10% per 10 seconds and reaching 100% over 80 seconds. Two parallel halt conditions operated: (i) a pause condition in which any subsequent epoch failing the ≥90% N3 criterion (transition to N1, N2, or REM) triggered an immediate pause, with volume reset to 20%, the N3-epoch counter reset to zero, and the playback position within the auditory file preserved—so that resumption (after re-accumulating three N3 epochs) continued from the prior position; and (ii) a full-stop condition in which detection of wakefulness (alpha activity / unprompted awakening) signalled by the technician via the RETURN key triggered immediate cessation, with volume reset to 20%, the N3-epoch counter reset to zero, and the playback position reset to the beginning of the auditory file. After either halt, restoration of stimulation required the same 90-second SWS-stable onset criterion. Cumulative sound delivery was capped at 600 seconds (10 minutes) per night. Pre-SES SUDs were measured during the waking sound exposure session in the evening prior to the overnight sleep session. Post-SES SUDs were measured the following morning by re-presenting the same trauma-associated sound during wakefulness. This pre-post comparison thus reflects the change in subjective distress to the identical auditory stimulus before and after a single night of SES.

Although cue selection required participants to briefly listen to candidate trauma-associated sounds during waking to provide SUDs ratings, this transient setup procedure is operationally distinct from the sustained, manualized therapeutic exposure that characterizes PE or EMDR. SES is therefore not intended to replicate or replace these evidence-based psychotherapies; rather, the waking cue-selection step is analogous to a baseline assessment procedure, with the therapeutic intent confined to the subsequent SWS-timed auditory delivery.

### Follow-Up (Visit 5)

One to four weeks post-intervention, follow-up assessments were conducted at participants’ regular medical facilities. Reassessments comprised the PCL-5 [66], PDS-5 [67], BDI-II [68], PSQI [69], SDS [70], and SUDs [71] obtained during a repeat waking sound-exposure session, together with adverse-event surveillance.

### Outcomes

Because this was the first study to utilize trauma-associated auditory cues for TMR in PTSD patients, the primary outcome was feasibility, expressed as a two-step funnel: (a) the proportion of patients who entered the sound-development phase with candidate personalized stimuli prepared and proceeded to overnight sleep-based stimulation (i.e., completed Visits 0–4 through to overnight stimulation entry); and (b) the proportion of patients who successfully completed the overnight sleep-based auditory stimulation procedure among those who entered Visit 4. The primary feasibility denominator for the second step is the n = 6 participants who proceeded to Visit 4; the first step denominator is the n = 8 participants who entered the sound-development phase with candidate personalized stimuli prepared (after exclusion of n = 5 prior to sound development due to ineligibility); a finalized SES stimulation sound was reached for n = 6 of these eight participants (H01, H02, H03, H08, H09, H14). Secondary exploratory outcomes included adverse events; pre-to-post changes in subjective and physiological fear responses (SUDs, skin conductance, heart rate); clinical symptoms; accuracy of sound presentation during SWS (operationalized as the percentage of cumulative sound-presentation time that overlapped with N3 sleep); sleep architecture changes; and any malfunctions in protocol implementation. The relative change formula for SUDs and skin conductance is provided in the Statistical Analysis section.

### Equipment and Data Handling

Home-based EEG was conducted using the portable “InSomnograf” system (S’UIMIN Inc.) with forehead and bilateral mastoid placements. During overnight SES, EEG and biometric data were collected using S’UIMIN’s advanced recording systems. Auditory cues were delivered via silicone earphones using encrypted Bluetooth. Skin conductance and pulse were measured with Empatica’s EmbracePlus. All personal data were anonymized prior to transfer and stored securely on encrypted platforms (AWS, Google, Jotform) in compliance with NCNP Ethics Committee protocols. Ethics monitoring and adverse event reporting followed approved procedures. Three device-related events were recorded during the overnight stimulation period in three of six participants. (i) In one participant, a programmed long-press of the event-tagging button on the sound-generator console did not register on the first three attempts; the on-call technician retried while standard white-noise playback continued, and tagging succeeded on the fourth attempt with time-stamping preserved. (ii) In another participant, after the SES sound stimulation had been completed, a power shortage on the EEG-recording PC terminated EEG capture during the latter portion of the night; protocol delivery and the participant’s completion of the intervention were unaffected. (iii) In a third participant, an incorrect earphone was attached, and a button activation before sleep onset triggered playback of its built-in rain sound; the earphone was replaced with the correct one and SES sound stimulation continued without further incident. None of these events caused arousal of the participant or premature termination of the protocol, and none affected the participant’s completion of the overnight SES intervention or eligibility for primary endpoint inclusion. Recurrence-prevention measures implemented after the trial included pre-procedure verification of the event-tagging button response on the sound- generator console, redundant power provisioning for the EEG-recording PC at the P1 Unit, and substitution with the correct earphone. All three participants were retained in the primary feasibility analysis.

### Blinding and Bias Control

Because auditory stimulation required real-time EEG monitoring with operator intervention, the sleep technicians who delivered stimulation could not be blinded to stimulation epochs. Post-hoc sleep staging and scoring of stimulus-time-locked events were performed without blinding to protocol version or stimulation timing, and formal inter-rater reliability (Cohen’s κ) was not assessed in this feasibility phase. Clinical outcome assessors administering follow-up PCL-5 and SUDs ratings were likewise not blinded to version assignment. These limitations are inherent to an open-label feasibility design and will be addressed in future adequately powered randomized trials through masked outcome assessment and independent blinded scoring.

### Statistical Analysis

Descriptive statistics were calculated for demographic variables and the number of stimulation sessions. As the primary outcome (second-step denominator), the proportion of participants who completed the overnight sound stimulation among those who proceeded to Visit 4 was calculated. The first-step proportion (proportion of participants reaching overnight stimulation among those who entered the sound-development phase with candidate personalized stimuli prepared) was tabulated separately and is reported in Figure 2 (CONSORT flow diagram). The frequencies of adverse events and device malfunctions were also summarized. To express the relative change in SUDs from the baseline value before stimulation, the SUDs change was defined as (post – pre) / pre × 100%, such that negative values indicate post-procedure reduction in distress.

Summary statistics for SUDs and other scores including PCL-5 and PDS-5 were calculated at each time point, and the changes in SUDs and other scores were also tabulated. As post-hoc exploratory analyses, one-sample t-tests were performed for each pre-post outcome (SUDs change, PCL-5 total/intrusion/avoidance/negative/hyperarousal, PDS-5) to examine whether the changes were significantly less than zero (i.e., indicating post-procedure improvement under the unified sign convention). In addition, two-sample t-tests and estimation of the mean and confidence interval (CI) with Satterthwaite approximation were conducted to compare the changes in scores between patients enrolled before (Version A) and after the protocol amendment (Version B). A two-sided significance level of 5% was used.

### Rationale for Parametric Inference in a Very Small, Unequal-Sample Feasibility Cohort

Given the feasibility framing and small, unequal sub-group sizes (Version A, n = 2; Version B, n = 4), our use of parametric inference warrants explicit justification. SUDs ratings often exhibit skewed distributions and recognised psychometric limitations, and strict normality cannot be assumed in small samples. However, rank-based alternatives such as the Mann–Whitney U test are mathematically incapable of producing a significant result under the present sample configuration: with n₁ = 2 and n₂ = 4, the total number of possible rank permutations is only C(6,2) = 15, so the minimum attainable two-sided p-value is 2/15 ≈ 0.133. This value cannot reach the conventional α = 0.05 irrespective of how large the true between-group separation might be, rendering non-parametric tests categorically unable to detect any exploratory signal under this design.

By contrast, large-scale simulation work has shown that at sample sizes as small as n = 2 per group, Student’s and Welch’s t-tests retain control of the Type I error at or below the nominal α under normal as well as markedly skewed distributions, leaving no principled objection to parametric inference in this setting [72]. We applied Welch–Satterthwaite-corrected t-tests. We further acknowledge that when both group sizes are below seven, the Welch–Satterthwaite approximation has been shown to under-represent uncertainty in certain variance configurations, yielding confidence intervals narrower than their nominal coverage [73]. This residual bias is, however, preferable to the categorical inability of rank-based alternatives to detect any effect at α = 0.05 under the present sample structure. Accordingly, all t-test p-values and confidence intervals reported here should be interpreted as measures of exploratory signal strength intended to calibrate effect size for the design of a subsequent adequately powered randomized controlled trial, rather than as confirmatory evidence of efficacy. No multiplicity correction was applied across the multiple exploratory outcomes (SUDs, PCL-5/PDS-5/BDI-II/PSQI/SDS, sleep architecture, HRV, SCL); all reported p-values are nominal and exploratory.

### Sensitivity Analyses: Statistical Robustness

Because protocol versions were implemented sequentially (Version A: September–October 2024; Version B: March–June 2025) rather than allocated randomly, version assignment and enrollment date are near-perfectly collinear in the present cohort (Pearson correlation between version and enrollment month = 0.96). Analysis of covariance (ANCOVA) including enrollment month as a covariate is therefore uninformative, as the version and time coefficients cannot be independently estimated. In lieu of ANCOVA, we conducted three sensitivity analyses. First, we enumerated all C(6,2) = 15 possible assignments of the six participants to Version A (n = 2) versus Version B (n = 4) and computed the mean between-version difference for each, yielding an exact permutation distribution. Second, we performed leave-one-out Welch t-tests, dropping each Version B participant in turn (dropping a Version A participant leaves n_A = 1, rendering group variance and the Welch t-test mathematically undefined). Third, we computed the Spearman correlation between each outcome and enrollment month as a version-agnostic test of temporal trend. Results are summarised in Supplementary Table S2 and interpreted in the Discussion.

## Author Contributions

M.S. and Y.Ki. conceptualized the study. K.I. served as the principal investigator and oversaw clinical research implementation, patient recruitment, and coordination across study sites. K.Z. designed and developed the auditory stimulus generation system. A.H. contributed to protocol development, patient referral, and stimulus creation. T.M. served as the responsible physician for the Phase 1 Unit at The University of Tokyo Hospital and facilitated overnight study sessions. M.T. conducted real-time sleep-based auditory stimulation, supervised the sleep technician team, and performed sleep scoring analyses. Y.S. served as the initial responsible physician at the University of Tokyo, facilitating site access, coordination with the Department of Neuropsychiatry, and patient referral. M.O. performed statistical analyses and contributed to writing the manuscript. Y.Y. implemented the sound playback program and contributed to stimulus creation. L.S. analyzed skin conductance response data, created figures, and contributed to writing the original draft. H.S. performed device and sound playback program validation, managed overnight sleep data quality, and assisted with protocol implementation. C.K. contributed to auditory stimulus data analysis, device validation, and protocol implementation. G.B. contributed to writing the original draft. I.W. supported overall research planning and implementation. H.Ki. contributed to device development. M.Y. contributed to device development and provided overall research support. Y.Ki. contributed to conceptualization, clinical research planning, implementation, and patient recruitment support. M.S. supervised the study, managed the coordinating center and data center, designed device specifications, and wrote the manuscript.

## Acknowledgements

This work was primarily conducted at the International Institute for Integrative Sleep Medicine (WPI-IIIS), University of Tsukuba. We thank Drs. K. Kuriyama, A. Kawamura, M. Oe, S. Lin, and Y. Toshishige for valuable suggestions on the study protocol; Dr. S. Hirohata for patient recruitment and valuable suggestions; Drs. E. Sakakibara and D. Koshiyama for on-call coordination at The University of Tokyo Hospital; Dr. M. Ohta and M. Yanagida for support at the Phase 1 Unit; Dr. T. Matsui for advice on skin conductance measurements; Dr. N. Kato, Dr. M. Sou, Dr. T. Machino, and Dr. T. Saito for valuable suggestions; W. Nakano and Y. Shimazu for clinical assessment support; S. Nakamura, C. Tanigawa, K. Shibasaki, and K. Fukuda (S’UIMIN Inc.) for technical support; K. Hashimoto, K. Maruo, T. Kokubo, and T. Mochizuki for research support; C. Kawasaki for research coordination; and I. Sekiguchi, M. Sakurai, and I. Kimura for administrative support.

This study was registered at the Japan Registry of Clinical Trials (jRCT1030230706) and approved by the Ethics Committee of the National Center of Neurology and Psychiatry (approval no. B2023-118). This work was supported by the Japan Agency for Medical Research and Development (AMED) (JP21zf0127005, JP21km0908001, JP23wm0525003 to M.S.), the Japan Society for the Promotion of Science (JSPS) (24H00894, 23H02784, 22H00469, 26H02428 to M.S.; 25KJ0664 to C.K.), and the Takeda Science Foundation (to M.S.).

## Data Availability

Source data underlying main and supplementary figures are provided as Source Data files, excluding direct personal identifiers (access subject to a Data Use Agreement and approval from the NCNP Ethics Committee, given the sensitive nature of trauma-related audio descriptions). Per-participant trauma-content metadata will not be disclosed publicly to minimize re-identification and re-traumatization risk; access for qualified researchers may be requested from the corresponding author.

## Code Availability

All statistical analyses reported in this study were conducted using standard scientific computing libraries with default parameters as described in Methods §Statistical Analysis and §Sensitivity Analyses; no custom analytical code beyond standard library function calls was required. Detailed parameter settings and analysis workflow are documented in the supplementary materials and are available from the corresponding author upon reasonable request. No proprietary or commercial code was used.

## Supplementary Figure Legends

**Figure S1.**
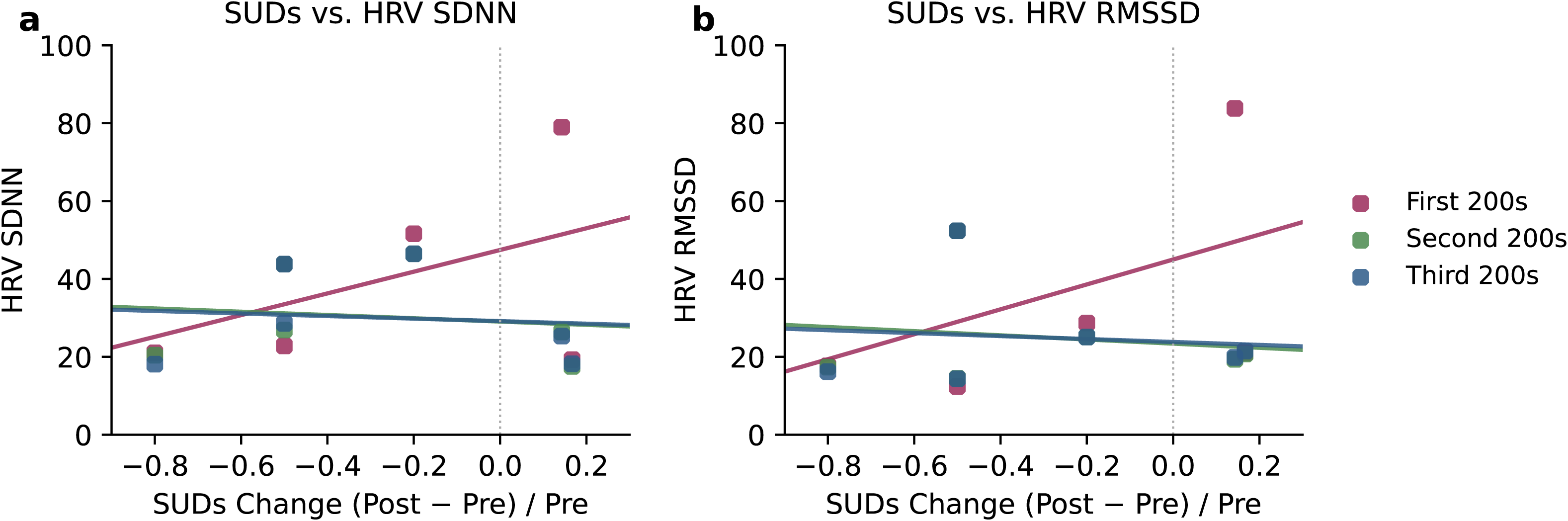
Association between change in SUDs and heart rate variability (HRV). Scatterplot showing the relationship between proportional change in SUDs, calculated as (Post − Pre) / Pre, and (a) HRV standard deviation of normal-to-normal intervals (SDNN) and (b) HRV root mean square of successive differences (RMSSD). Data are shown for the first 200 s (maroon), second 200 s (green), and third 200 s (blue) epochs. Lines represent linear regression fits for each epoch.

**Figure S2.**
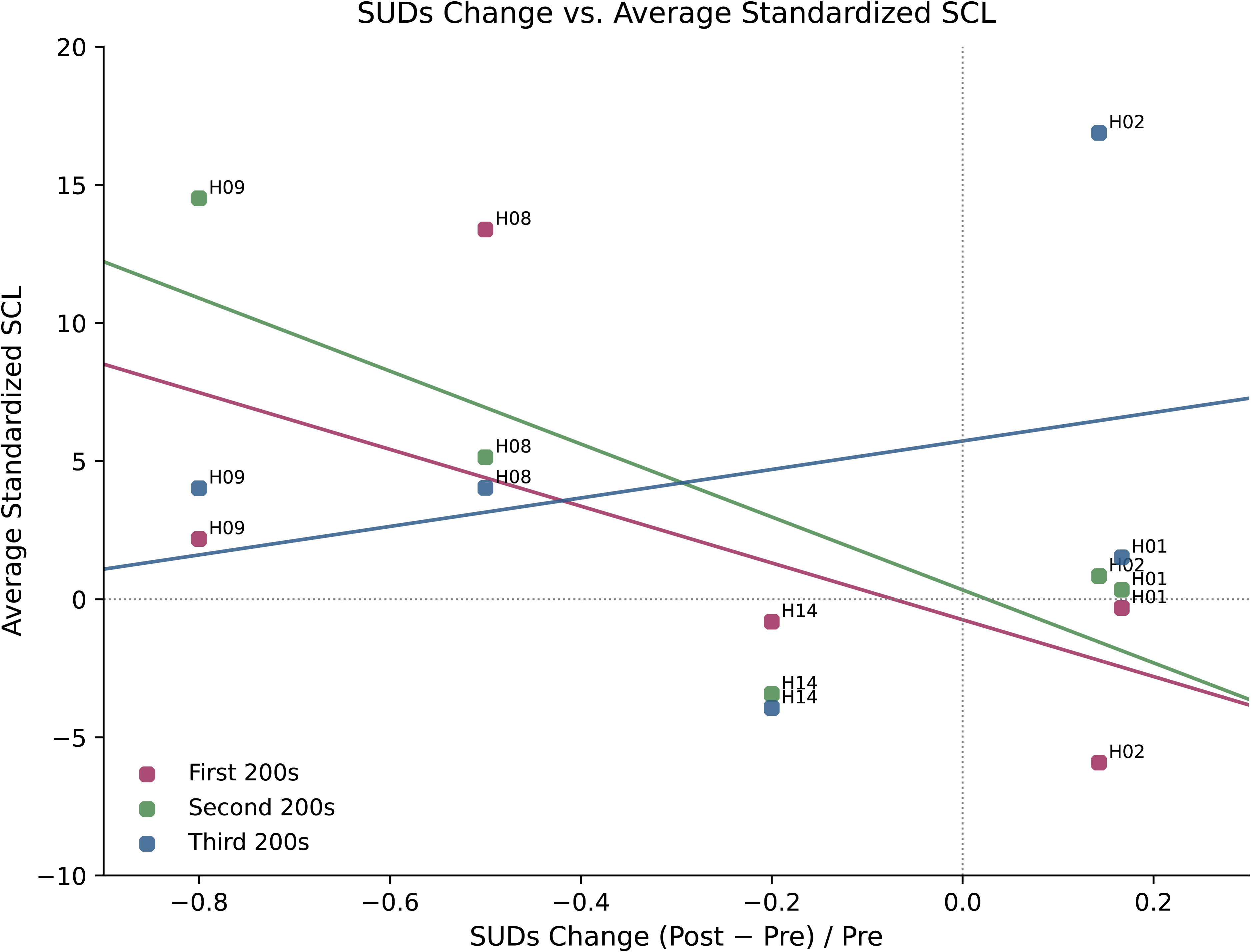
Association between change in SUDs and skin conductance level (SCL). Scatterplot showing the relationship between proportional change in SUDs, calculated as (Post − Pre) / Pre, and average standardized skin conductance level (SCL) (n = 5; H03 was excluded due to SCL recording failure). SCL values were standardized within participants using the 90 s pre-epoch baseline (z = (SCL − baseline mean) / baseline SD). Data are shown separately for the first 200 s (maroon), second 200 s (green), and third 200 s (blue) epochs. Lines represent linear regression fits for each epoch.

**Figure S3.**
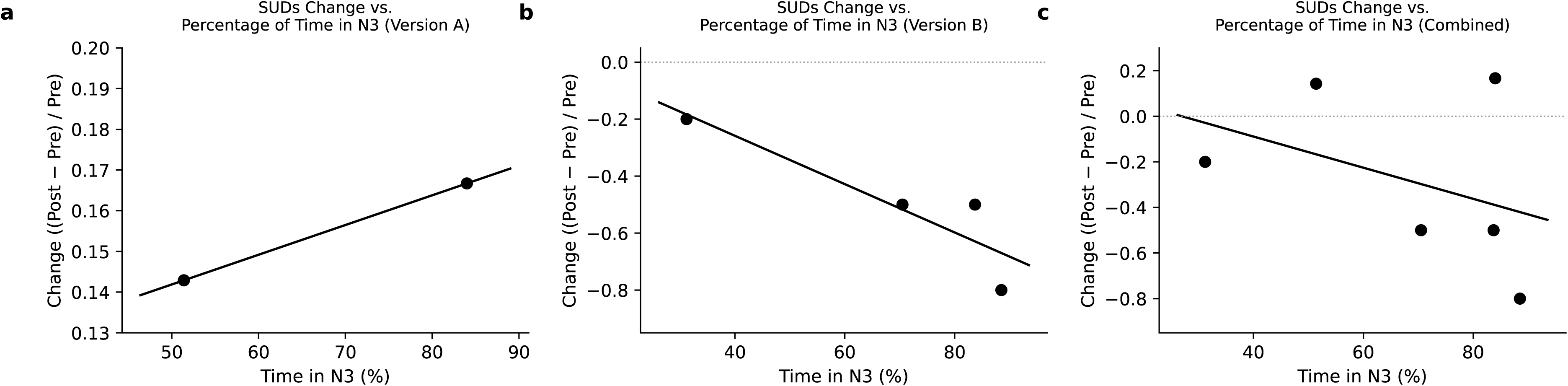
Association between change in SUDs and percentage of time spent in N3 sleep. Scatterplots showing the relationship between proportional change in SUDs, calculated as (Post − Pre) / Pre, and percentage of time spent in N3 sleep during SES in participants for (a) Version A, (b) Version B, and (c) the combined sample. Individual dots represent a participant. Lines represent linear regression fits.

**Figure S4.**
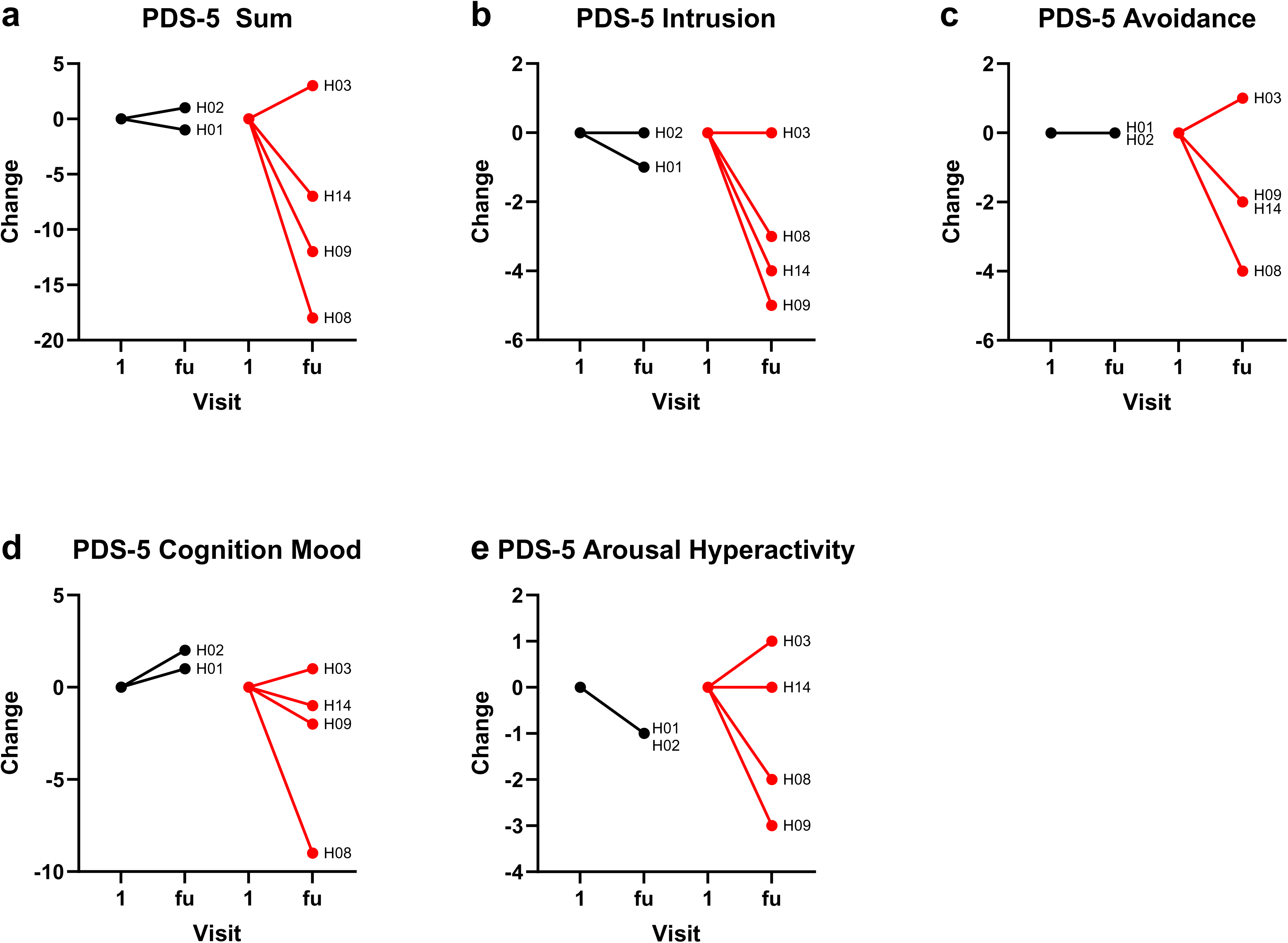
Visit 1 to follow-up (fu) changes in PDS-5 sub-scores. Individual participant data are shown for (a) PDS-5 Sum, (b) PDS-5 Intrusion, (c) PDS-5 Avoidance, (d) PDS-5 Cognition Mood (changes in cognition and mood), and (e) PDS-5 Arousal Hyperactivity (arousal and hyperactivity) sub-scores obtained at visit 1 and at follow-up. Participants receiving Version A are denoted in black and those receiving Version B in red.

**Figure S5.**
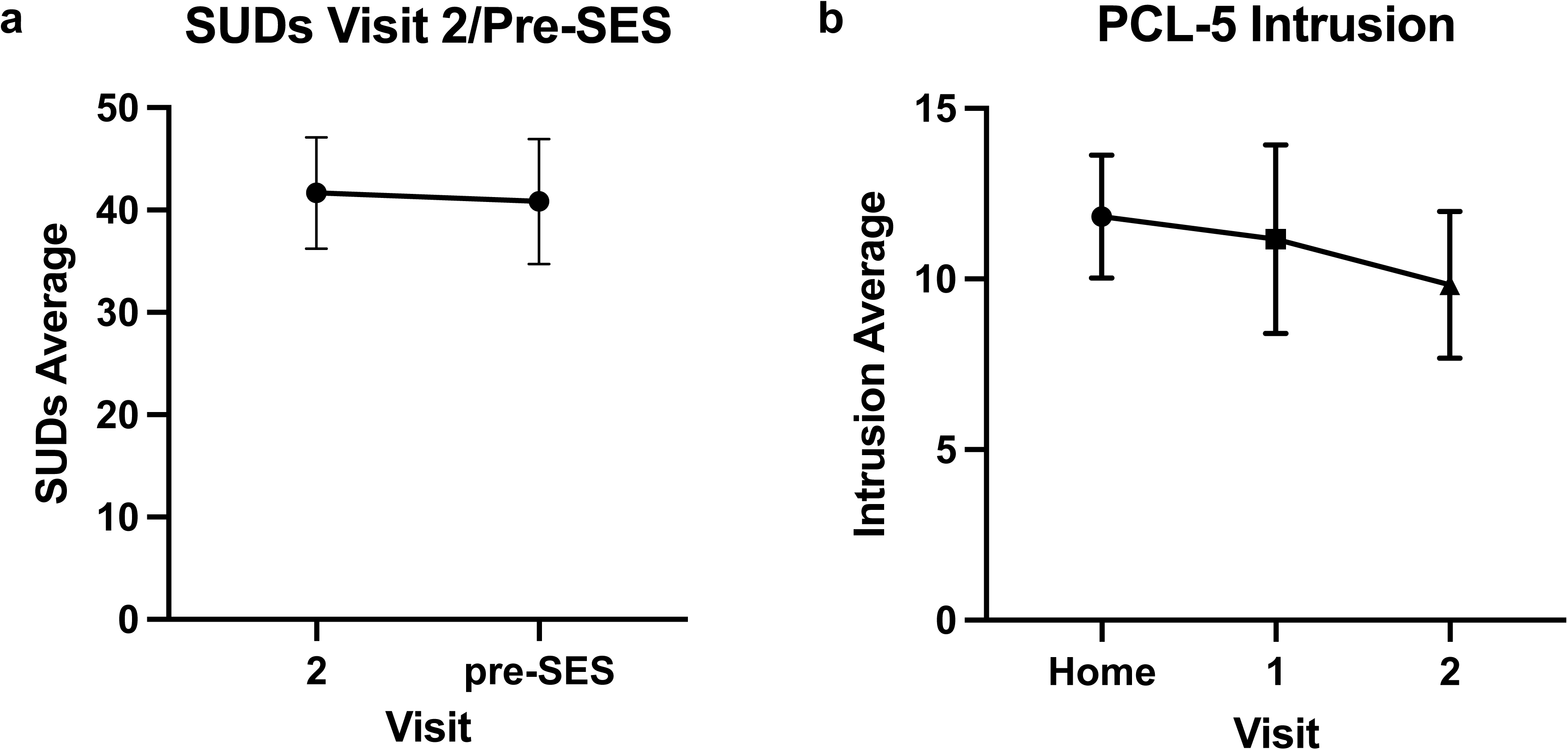
Raw SUDs and PCL-5 scores across study visits before SES. Group means ± standard error of the mean (SEM) are shown for (a) SUDs rating comparison between visit 2 and pre-SES and (b) PCL-5 intrusion score comparison between the home screening, visit 1, and visit 2.

**Figure S6.**
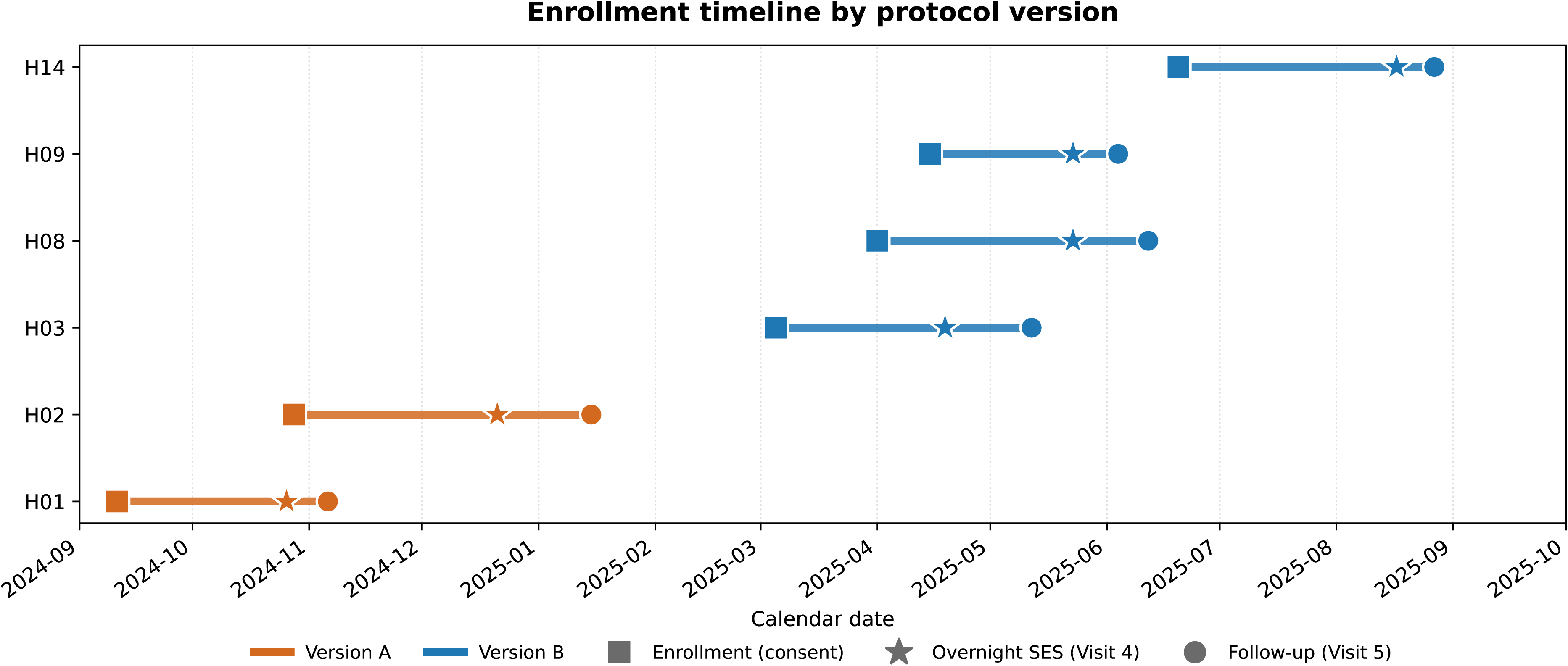
Enrollment timeline by protocol version. Calendar date plot showing the enrollment, overnight SES (Visit 4), and follow-up (Visit 5) timepoints for each of the six participants, color-coded by protocol version. Visualizes the sequential implementation of the two protocol versions, leaving a four-month gap between the last Version A and the first Version B enrollment.

