## Supplementary Table S1 for "Sound Exposure During Sleep (SES) in PTSD Patients: An Open-Label Feasibility Study"

**Supplementary Table S1. Concomitant medications at enrollment, by participant.**

*Concomitant medication status for each of the six participants who completed the overnight SES procedure (Version A: H01, H02; Version B: H03, H08, H09, H14). All concomitant medications recorded in the enrollment Case Report Form (CRF) are listed. Drug names are shown at the INN (generic) level; commercial names are retained for over-the-counter or multi-ingredient preparations where INN coding is not applicable. Dashes (—) indicate no medication in that class at enrollment. The bottom row shows class-level counts across the six participants.*

| **Patient ID** | **Protocol version** | **Antidepressant (incl. SSRI)** | **Anxiolytic-class drug listed** | **Hypnotic** | **Antipsychotic** | **Mood stabilizer** | **Other (non-psychotropic)** |
| --- | --- | --- | --- | --- | --- | --- | --- |
| **H01** | A | — | Clonazepam | — | Quetiapine fumarate | Valproate sodium | Memantine hydrochloride‡  Bromocriptine mesilate  Pramipexole hydrochloride  Omeprazole sodium  Loxoprofen sodium hydrate  Rebamipide  Bifidobacterium preparation |
| **H02** | A | — | — | Lemborexant | — | — | Semaglutide  Bifidobacterium preparation  Rosuvastatin calcium  Mirogabalin besilate  Tramadol hydrochloride  Metoclopramide  Magnesium oxide  Tocopherol acetate  Local anesthetic (for low back pain) |
| **H03** | B | — | — | — | — | — | Levothyroxine sodium hydrate  Bilastine  Betamethasone / d-chlorpheniramine maleate  Valaciclovir hydrochloride |
| **H08** | B | Escitalopram oxalate [SSRI] | Clotiazepam | Lemborexant  Eszopiclone | Levomepromazine maleate  Brexpiprazole | — | Mosapride citrate hydrate  Kami-shoyo-san extract  Memantine hydrochloride‡ |
| **H09** | B | Sertraline hydrochloride [SSRI]  Trazodone hydrochloride | Bromazepam  Clonazepam | Eszopiclone | Risperidone (oral tablet)  Risperidone (oral solution) | — | Magnesium oxide  Acetaminophen  d-Chlorpheniramine maleate  Montelukast sodium  Levocetirizine hydrochloride |
| **H14** | B | — | — | — | — | — | Memantine hydrochloride‡  Levonorgestrel / ethinylestradiol  Ohta's gastric digestant A  Loxoprofen sodium hydrate  Aneron Niscap  Ibuprofen  Tranexamic acid  Dried aluminum hydroxide gel  Kakkon-to extract  Yokukansan extract |
| **n=6** | **A:2 / B:4** | **2 (33.3%); 2 SSRI** | **3 (50.0%)*** | **3 (50.0%)** | **3 (50.0%)** | **1 (16.7%)** | **All 6 (100%) on ≥1 non-psychotropic** |

*Footnote. *The 'Anxiolytic-class drug listed' column counts any benzodiazepine or GABAergic anxiolytic regardless of indication (3 / 50.0%); this includes clonazepam in H01, which was clinically prescribed together with dopamine agonists (bromocriptine, pramipexole) most plausibly for restless-legs/sleep-related indications rather than as a primary anxiolytic. By contrast, the main-text Table 1 column 'Anxiolytics' uses an indication-based classification and therefore reports 2 (33.3%); the difference between the two counts reflects only this difference in classification basis, not a data discrepancy.*

*Abbreviations. SSRI, selective serotonin reuptake inhibitor; INN, International Nonproprietary Name. ‡Memantine (an N-methyl-D-aspartate receptor antagonist) was taken as stable background therapy by H01, H08, and H14 (n = 3); doses and regimens were unchanged throughout the SES protocol and follow-up period. A descriptive sensitivity analysis comparing exploratory symptom outcomes by memantine status is reported in Supplementary Table S3.*

*Data source. Concomitant medication CRF entries coded against PMDA generic-name (INN) database; class assignments (antidepressant / anxiolytic / hypnotic / antipsychotic / mood-stabilizer / other) were made by drug-mechanism lookup against PMDA drug information. Master-coding spreadsheet and signed PDF copy are retained on the study record.*
