## Supplementary Table S2 for "Sound Exposure During Sleep (SES) in PTSD Patients: An Open-Label Feasibility Study"

**Supplementary Table S2. Sensitivity analyses assessing the statistical robustness of the observed between-version difference under temporal confounding (Version A: n = 2, enrolled Sep–Oct 2024; Version B: n = 4, enrolled Mar–Jun 2025).** Because version assignment and enrollment date are near-perfectly collinear in this cohort (Pearson r = 0.96), these analyses do not disentangle version from calendar time; the temporal-confounding limitation is addressed in the Limitations section of the Discussion.

| Outcome | Welch diff (B − A) | Welch 95% CI | Welch p | Perm rank / 15 | Perm p (exact, two-sided) | LOO diff range (B drops)† | LOO p range (B drops)† | Spearman r (vs enrollment month) | Spearman p | Within-B Spearman r (n = 4)† | Within-B Spearman p† |
| --- | --- | --- | --- | --- | --- | --- | --- | --- | --- | --- | --- |
| PCL-5 intrusion change | −7.00 | [−11.61, −2.39] | 0.015 | 1 / 15 | 0.067 | −8.00 to −6.33 | 0.015 – 0.037 | −0.647 | 0.165 | +0.333 | 0.667 |
| PCL-5 total change | −14.00 | [−30.42, 2.42] | 0.073 | 2 / 15 | 0.133 | −17.17 to −9.83 | 0.097 – 0.217 | −0.754 | 0.084 | −0.316 | 0.684 |
| SUDs change (%) | −65.48 | [−104.24, −26.72] | 0.012 | 1 / 15 | 0.067 | −75.48 to −55.48 | 0.016 – 0.063 | −0.603 | 0.205 | +0.500 | 0.500 |

**Notes.** Welch diff (B − A), between-version mean difference estimated by the Welch t-test with Satterthwaite-approximated degrees of freedom. Welch 95% CI, two-sided 95% confidence interval for the between-version mean difference. Perm rank / 15, the rank of the observed absolute between-version difference among all C(6,2) = 15 possible assignments of the six participants to Version A (n = 2) versus Version B (n = 4); Perm p, exact two-sided p-value computed as #{|diff_perm| ≥ |diff_obs|} / 15. The combinatorial floor of this exact distribution is 1/15 ≈ 0.067 when the observed split yields the single most extreme absolute difference in the enumerated set. LOO = leave-one-out Welch t-test. Spearman r and p are computed between each outcome and enrollment month, coded ordinally 1 = Sep 2024 through 10 = Jun 2025 (H01 = 1, H02 = 2, H03 = 7, H08 = 8, H09 = 8, H14 = 10; n = 6); this provides a version-agnostic test of temporal trend. Within-Version-B Spearman r (n = 4), Spearman correlation between outcome and enrollment month computed within Version B participants only (H03, H08, H09, H14) — an additional version-agnostic check; non-significance and inconsistent direction across outcomes argue against a within-Version-B calendar-time trend.

† Leave-one-out Welch testing was restricted to Version B participants because dropping a Version A participant leaves n_A = 1, rendering the group variance and the Welch t-test mathematically undefined. The absolute between-version mean differences remained highly stable under descriptive A-drops as well: PCL-5 intrusion change −6.00 (drop H01) to −8.00 (drop H02); PCL-5 total change −14.50 (drop H01) to −13.50 (drop H02); SUDs change −64.29 % (drop H01) to −66.67 % (drop H02). These A-drop descriptive estimates confirm the observed effect magnitude is not driven by either Version A participant individually. Sign convention. PCL-5 intrusion change and PCL-5 total change rows use the raw (FU − Pre) convention, where negative values indicate symptom improvement and a negative Spearman r against enrollment month indicates that later-enrolled participants showed larger improvements. The SUDs change (%) row now uses the same (Post − Pre)/Pre × 100 convention as the PCL-5 rows, where negative values indicate post-procedure reduction in distress and a negative Spearman r against enrollment month indicates that later-enrolled participants showed larger reductions. All rows in this table therefore share a unified orientation (negative = improvement), aligning the manuscript with Figure 4 and the source-data calculation pipeline.

**Source data and code provenance.** Exploratory symptom-outcome values were extracted from the Source Data file (sheets Fig4_SUDs_PrePost and Fig5_PCL5_Change). Sensitivity analyses were performed using an exhaustive enumeration of C(6,2) = 15 splits for the permutation test, Welch's t-test (unequal variances) and Spearman's rank correlation for the time-trend correlation. Enrollment months were coded by the calendar month of consent. Independent re-execution of the analyses by a second computational pass reproduced all reported values to within rounding (≤ 0.001 in p, ≤ 0.01 in r and effect sizes).
