## Supplementary Table S3 for "Sound Exposure During Sleep (SES) in PTSD Patients: An Open-Label Feasibility Study"

**Supplementary Table S3. Descriptive sensitivity analysis of exploratory symptom outcomes by memantine status.** Three of six participants (H01, H08, H14) were on stable memantine therapy at enrollment, with doses unchanged throughout the trial period; the remaining three participants (H02, H03, H09) were not. Pre-treatment baselines, post-treatment values, and change scores are presented descriptively for both subgroups. Given the very small subgroup sizes (n = 3 each), formal hypothesis testing is mathematically underpowered—the minimum two-sided p-value attainable by exact permutation enumeration over all C(6,3) = 20 group splits is 0.10—and is not reported.

**Per-participant outcomes (with baselines)**

| Patient | Version | Memantine | PCL-5 sum baseline | PCL-5 sum change | PCL-5 intrusion change | SUDs baseline | SUDs change (%) |
| --- | --- | --- | --- | --- | --- | --- | --- |
| H01 | A | + | 44 | 0 | +3 | 30 | +16.67 |
| H02 | A | − | 61 | +1 | +1 | 35 | +14.29 |
| H03 | B | − | 37 | −4 | −4 | 60 | −50.00 |
| H08 | B | + | 46 | −26 | −7 | 20 | −50.00 |
| H09 | B | − | 51 | −18 | −7 | 50 | −80.00 |
| H14 | B | + | 21 | −6 | −2 | 50 | −20.00 |

**Subgroup descriptive summary (means)**

| Subgroup | PCL-5 sum baseline (mean) | PCL-5 sum change (mean) | PCL-5 intrusion change (mean) | SUDs baseline (mean) | SUDs change (mean, %) |
| --- | --- | --- | --- | --- | --- |
| Memantine + (n=3) | 37.00 | −10.67 | −2.00 | 33.33 | −17.78 |
| Memantine − (n=3) | 49.67 | −7.00 | −3.33 | 48.33 | −38.57 |
| Difference (+ minus −) | −12.67 | −3.67 | +1.33 | −15.00 | +20.79 |

**Notes.** Mem+, on stable memantine therapy at enrollment (no dose change during trial); Mem−, not on memantine. Values are reported descriptively as means; no inferential statistics are presented because at n = 3 versus n = 3 the minimum two-sided p-value attainable by exact permutation is 0.10, rendering null-hypothesis significance testing structurally unable to detect even maximal between-group differences. The two subgroups differ at baseline: Mem+ participants exhibited lower mean baseline PCL-5 sum (37.0 vs. 49.7) and lower mean baseline SUDs (33.3 vs. 48.3) than Mem− participants. Reported change scores should therefore be interpreted in light of these baseline asymmetries (under the unified (post − pre)/pre × 100 sign convention, negative SUDs change values indicate post-procedure reduction in distress) (e.g., potential floor effects in the Mem+ subgroup) and the substantive mechanistic confound discussed in the main-text Limitations. Results are reported here for transparency, not as evidence of presence or absence of a memantine effect.

**Source data.** Per-participant outcomes and baselines extracted from the Source Data file (sheets Fig4_SUDs_PrePost and Fig5_PCL5_Change). Memantine status determined from the Concomitant Medication CRF (see Supplementary Table S1).
